# A new method to measure Baroreflex sensitivity impairment in Long Covid patients with Hyperadrenergic POTS-like symptoms

**DOI:** 10.1101/2025.08.05.25332555

**Authors:** Alejandro Sáinz-Jiménez, Ignacio Romero Fragoso, Guadalupe E. Salazar Calderón, Santiago Martínez Falcón, Hannah M. Luna, Jesus Portocarrero Nieto, Felipe Gonzalez-Alvarez, Tania Reyes-Cruz, Brayans Becerra-Luna, Raul Martinez-Memije, Erwin Chiquete, Karla M. Tamez-Torres, José Sifuentes-Osornio, Ruben Fossion, Bruno Estañol, José J. Aceves Buendía

## Abstract

Long Covid is the expression used to describe a variety of COVID-19 side effects lasting longer than three months. Within these side effects are cardiovascular alterations, such as Postural Orthostatic Tachycardia Syndrome (POTS) caused by an autonomic nervous system dysfunction.

A higher incidence of POTS and decreased baroreceptor sensitivity was found in Long Covid patients. Many of these patients suffer from orthostatic intolerance similar to those of POTS patients that do not strictly coincide with the criteria that have previously been established for POTS subtypes.

Therefore, to further describe the Orthostatic Intolerance we created a new method that better characterizes cardiovascular changes. Because these changes are reflected in the baroreceptor sensitivity measure, this new method consists in a geometrical analysis, where a decrease in baroreceptor sensitivity (BRS) is shown; this BRS measurement is added to the variables that have been considered, to improve the diagnostic of patients with POTS.

The proposed method does not necessarily require an algorithm like other methods in order to be performed, and generates a graph that can be easily interpreted. Furthermore, it gives information that aligns with the patient’s current heart rate variability status.

## 1. Introduction

Following the SARS-CoV-2 pandemic, patients who survived an acute COVID-19 infection presented with a wide variety of sequelae after 1-6 months of the acute phase of the disease. This set of symptoms is commonly referred to as Long Covid.

Among these sequelae, dysautonomia and orthostatic intolerance could be identified. Alongside several cases of Postural Tachycardia Syndrome (POTS) had a significant incidence rise from 1.42/1,000,000 to 20.3/1,000,000 person-year cases (Dulal et al., 2025), where 9 to 61% of infected people reported symptoms similar to those of POTS patients (Ormiston et al., 2022; Lee et al., 2024).

POTS-like symptoms have been previously identified after an acute viral infection (Grubb, 2008; Hickie et al., 2006) that could be associated with an abnormal inflammatory response (Medow & Stewart, 2007; Stewart, 2004; Low et al., 1995).

These POTS-like symptoms were referred to as orthostatic intolerance (OI), a broad term assigned to a group of heterogeneous pathologies with similar features like changes in heart rate and arterial blood pressure during the upright position (Ruzeih & Grubb, 2018; Gilmore et al., 2021; Pickering, 2003).

Some of the symptoms that are present within OI are: syncope, blood pooling in the lower limbs, an altered response of the baroreflex sensitivity with a change in the sympathetic nervous response; this results in several discomfort symptoms such as dizziness, blurry vision, fatigue and palpitations. All these appear or worsen with the upright position, and are relieved or diminished by returning to the supine position (Gilmore et al., 2021).

Within the pathologies described in OI, we can find POTS, neurocardiac syncope, orthostatic hypotension, vasovagal syncope, immediate orthostatic hypotension, cardioinhibitory syncope, among others (Gilmore et al., 2021).

Nevertheless, POTS-like symptoms and their intensity need to be reevaluated in order to establish a classification range for patients who do not reach the criteria for POTS, specifically the hyperadrenergic POTS subtype, characterized by an excessive tachycardia equal or greater than 30 bpm and a rise in systolic blood pressure (SBP) equal or greater than 10mmHg during the first 10 minutes of standing (Ruzieh & Grubb, 2018).

Some authors like Low et al., 2013 and Hira et al., 2025 show a change in baroreflex sensitivity (BRS) during active orthostatism test in POTS patients. Thus, we will use the BRS and heart rate variability (HRV) to segregate and define the subtypes of mild hyperadrenergic POTS.

## 2. Methods

### 2.1. Recruitment strategies for study participants

Patients were selected from a database containing those who had been infected with COVID-19 and presented symptoms after four weeks of the acute state of infection (Nalbandian et al., 2021).

Patients were first screened using the Compass 31 questionnaire, alongside a modified version of the same questionnaire (Compass 31+) designed to screen for concordance of COVID-19 sequelae with Long Covid; then a control group, which had no comorbidities such as diabetes, high blood pressure or other conditions, was selected (n=14).

Once these patients were selected, they underwent an active orthostatism test, where their blood pressure and heart rate were measured using a Finapress finometer with a finger sensor used for continuously monitoring and collecting hemodynamic data generated beat-by-beat.

The following protocol was done for all patients: the device was placed on their finger and then they were asked to lie in the supine position for 7 minutes. After this time, the patients were asked to stand up by themselves and stay that way for 7 minutes, breathing normally and trying not to move. Subsequently, they were asked to return to the supine position and remained so for another 7 minutes.

### 2.2. Stable state analysis

Once the Systolic blood pressure (SBP), Heart rate(Hr) and InterBeat Interval (IBI) were obtained, statistical analysis was performed.

All patient data was analyzed following the same procedure. Firstly, we selected the data periods corresponding to the steady states of SBP, HR and IBI values during the supine position and orthostatism. These stable periods were established when the patients were not moving and there were no external perturbations of the system; once this was done, we calculated the delta(Δ) value between the mean measurement of each position, and additionally, the mean and standard deviation were calculated for the previously mentioned variables.

To quantify Heart Rate Variability (HRV) we used Kubious software to calculate the linear variables: pNN50, SDNN and RMSSD values, and also to calculate the non linear **α**2 from the DFA (detrended fluctuation analysis) for each steady state.

In addition, we used a Poincaré graph with a 95% confidence level for the IBI with the aim to observe the beat by beat regularity and dispersion, as well as to calculate the Sd1 and Sd2 values in order to measure eccentricity for the adjusted orbit.

### 2.3. BRS Analysis

To measure the baroreflex sensibility (BRS), we developed an innovative graphical methodology based on the ΔIBI and ΔSBP, as described by Swenne et al. 2012, who define BRS as the change in InterBeat Interval in milliseconds per unit of change in Blood Pressure.

To calculate the baroreflex sensibility (BRS) using this graphical method, we took the data corresponding to previously selected SBP and Inter Beat Interval (IBI) steady states. Once selected, the differences between successive points were calculated to obtain values representing the pressure and heart rate changes per beat (ΔSBP & ΔIBI).

After this, we placed both Delta(Δ) columns on a scatter plot with ΔSBP on the X axis and ΔIBI on the Y axis; then an ellipse is adjusted over the data with 95% confidence to observe its behavior.

Following adjustment, we extracted the ellipse equation in its canonic form:

*Ax*^2^ + *Bxy* + *Cy*^2^ + *Dx* + *Ey* + *F* = 0. Once this was done, we normalized the adjusted ellipse to calculate its rotation angle against the X Axis, using a modified version of the standard rotation axis formula for conic equations to obtain what we refer to as the patient’s “Theta Angle”[°] (Fig. 1).

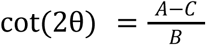

**Fig 1.**
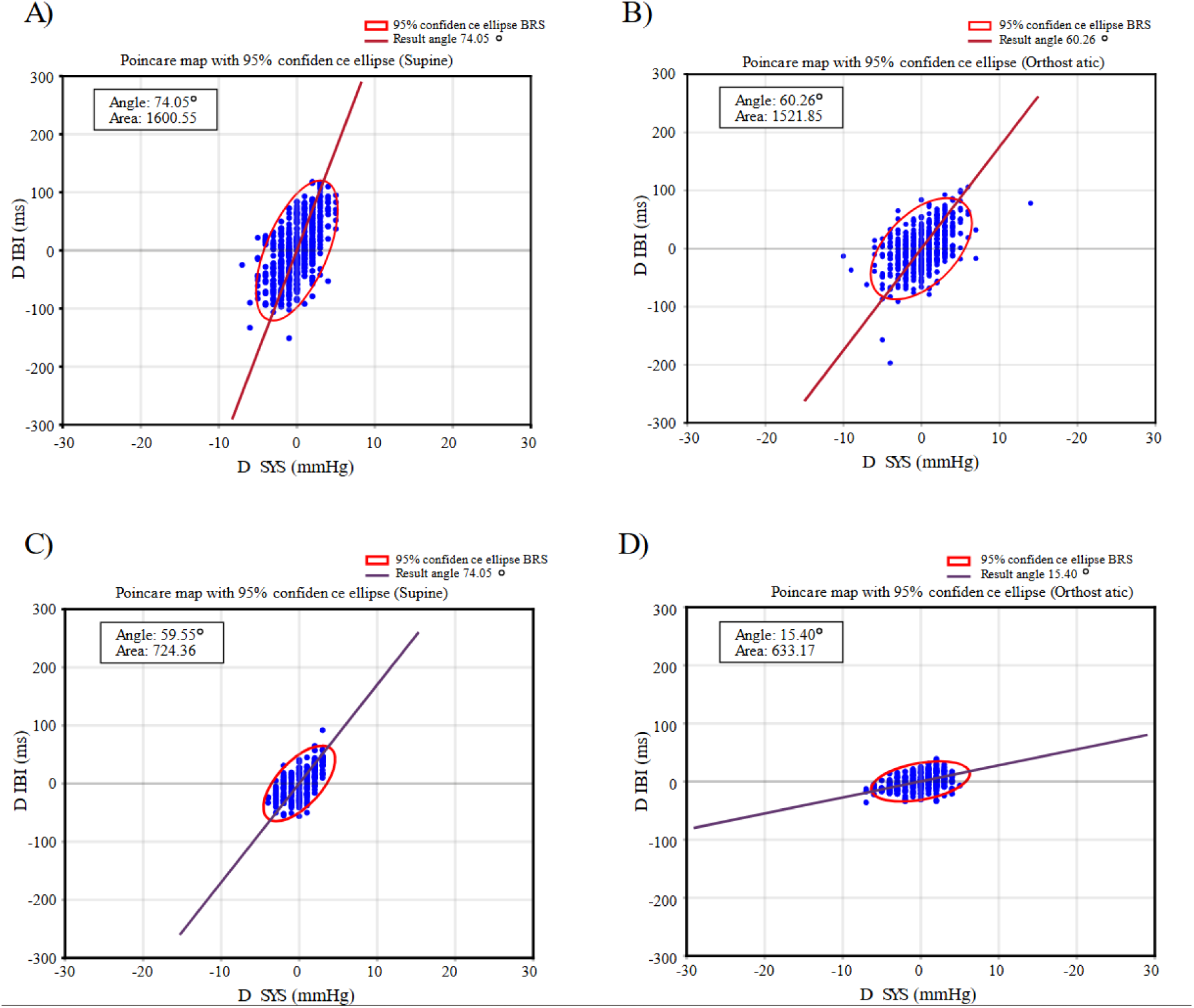
The Theta graphics corresponding to the supine(A) and orthostatic(B) positions of a healthy subject is shown. We show the scatter plot of the ΔSBP and ΔIBI values in blue, over it a 95% confidence ellipse was adjusted and is shown in red; additionally, a dark red line with the calculated Theta[θ] angle was overimposed. The axis scales were normalized to observe the natural response without modifying the raw delta(Δ) values. 1A) Theta graph during the supine period for a control subject. 1B) Theta graph during the orthostatism period for a control subject. 1C) Theta graph during the supine period for a pathological subject. 1D) Theta graph during the orthostatism period for a pathological subject.

From where we clear the θ variable to obtain:

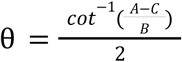

In order to normalize our data, we carried out the following procedure to ensure that this adjusted ellipse is always equal. We observed that the angle formed by normalizing the ΔSBP and ΔIBI columns is equal to the angle calculated when we rescaled the values of the canonical equation of the ellipse, where we multiply the values in the following way: A by 1, B by 10 and C by 100.

To validate this newly proposed method, we measured the patient’s BRS values using the sequence method (Bertinieri et al., 1985; Martínez-García et al., 2012). Furthermore, we calculated the BRS sensitivity using the difference between successive IBI values divided by the difference of successive SBP values, for which we also calculated their dispersion values, such as the mean, standard deviation and kurtosis (Fig. 2). Based on these values, we consider that we could also segregate the pathological groups suffering from hyperadrenergic POTS-Like Orthostatic Intolerance.

**Fig 2.**
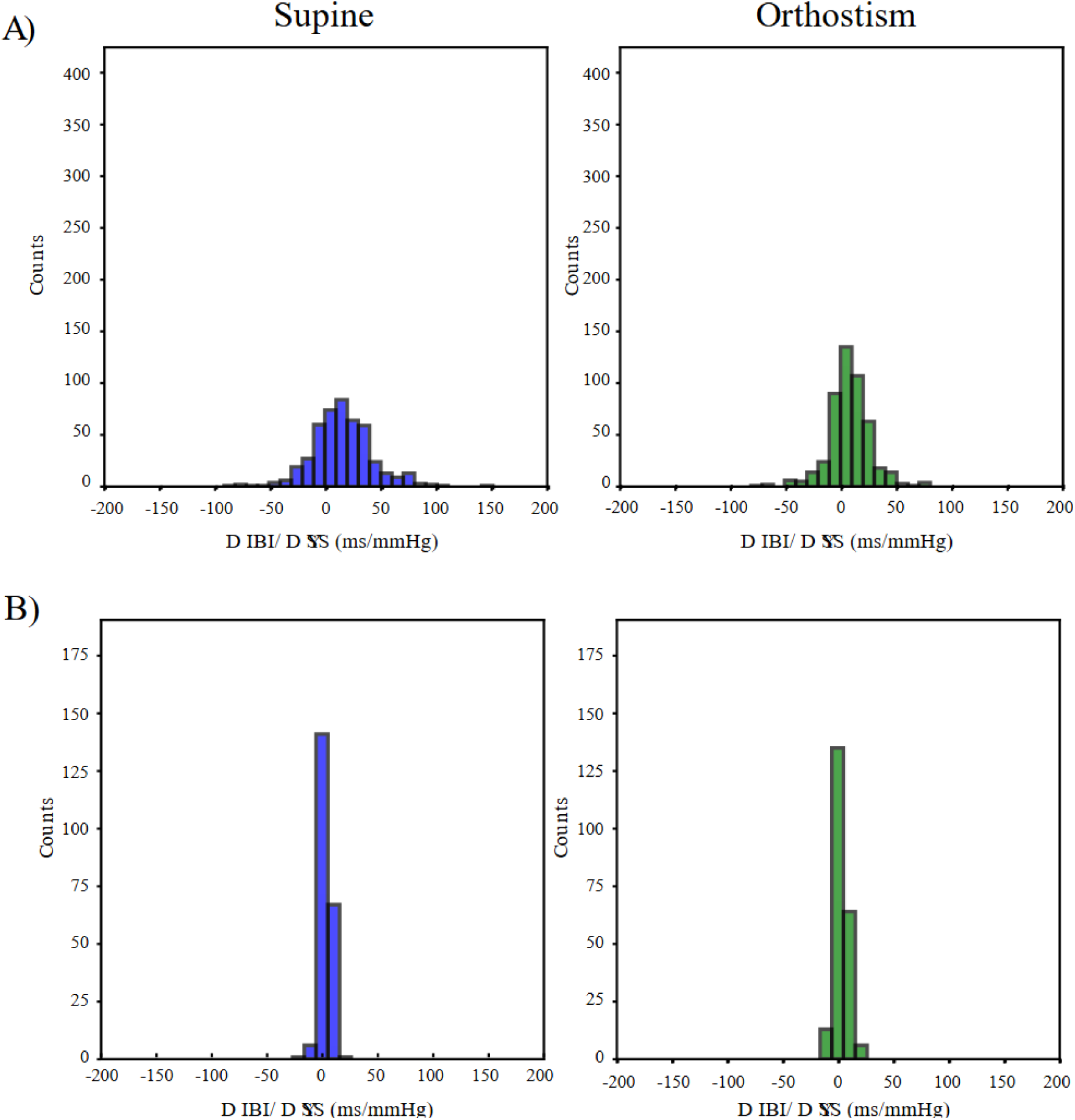
Histogram for the BRS values corresponding to the supine (left) and orthostatic (right) positions, obtained from the division of ΔIBI/ΔSYS beat-to-beat measurements. 2A) Histogram of the BRS values for a control subject during the supine and upright positions. 2B) Histogram of the BRS values for a pathological subject during the supine and upright positions.

Results will be reported as mean±standard deviation, for significance we performed the Mann-Whitney test. A two-tailed p-value of <0.05 was deemed to be statistically significant.

## 3. Results

We performed for all patients a modified version of the Compass 31 test and we found an increase in symptoms associated with the sympathetic system (45.55128±24.94489).

### 3.1. Long Covid patients show different types of responses during an active orthostatism test

Therefore, the changes in blood pressure and frequency in two positions (supine and orthostatism) were evaluated with the Finapress, and it was found that none of the patients met the criteria for hyperadrenergic POTS. Nevertheless, there is a wide distribution in blood pressure and heart rate delta values (13.72±12.66 mmHg, p=0.00504; 13.62±6.67 bpm, p=0.74684), with the corresponding coefficient of variation values SBP: 0.92338, HR: 0.48981 (Fig. 3A, B). Thus, the patients do not constitute a homogeneous group.

**Fig 3.**
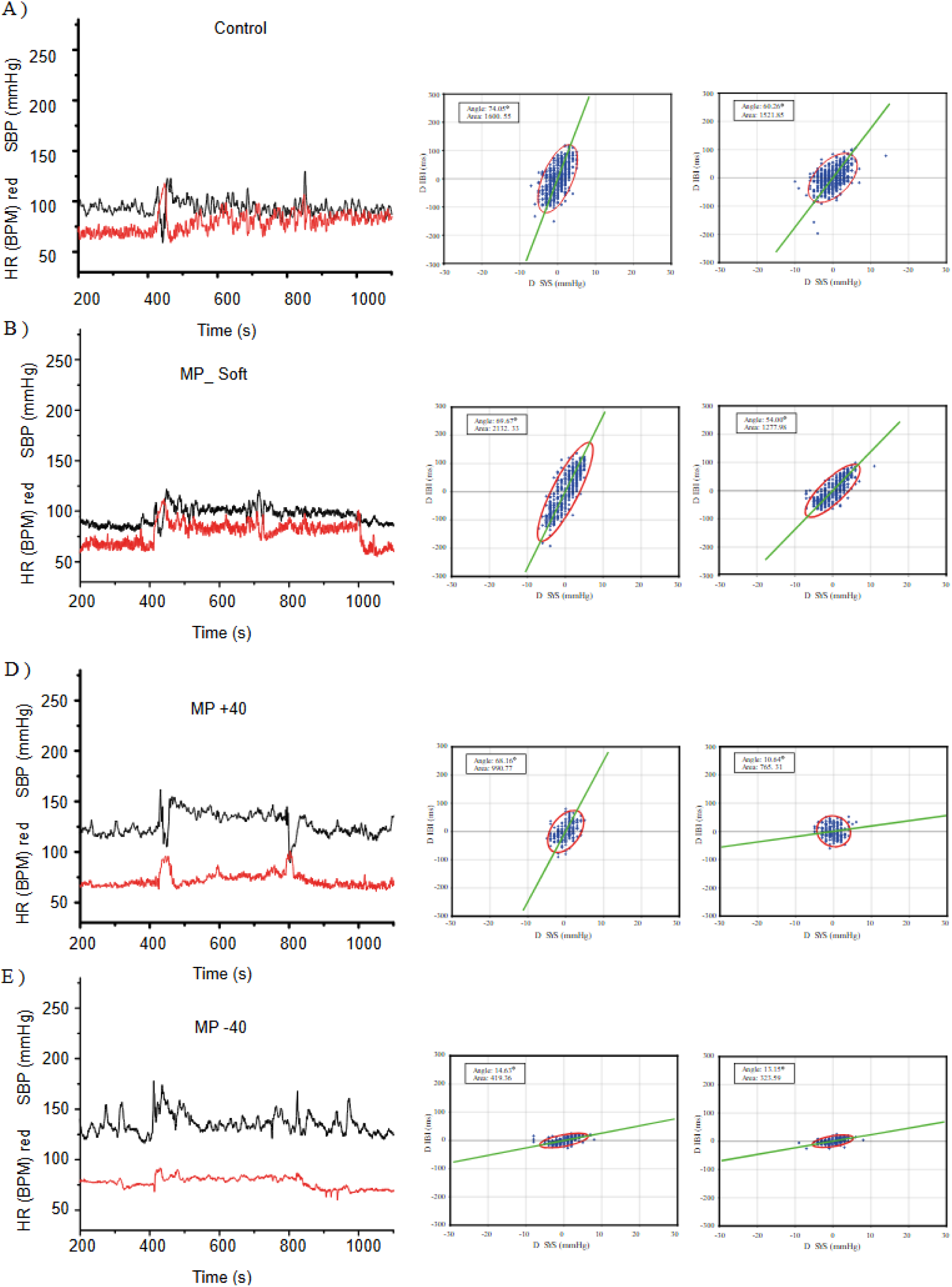
Measured SBP and HR values (colored in black and red, respectively) of one patient from each of our selected groups, along with the BRS response represented by the Theta angle for the supine and upright positions. 3A) SBP and HR values of a healthy control accompanied by their respective Theta graphs. 3B) SBP and HR values accompanied by their respective Theta graphs of a patient with Mild POTS and a soft change in the BRS response. 3C) SBP and HR values accompanied by their respective Theta graphs of a patient with Mild POTS and a change in angle >40° for the BRS response. 3D) SBP and HR values accompanied by their respective Theta graphs of a patient with Mild POTS and an attenuated BRS response.

### 3.2. BRS determined subgroups

Consequently, we decided to measure the BRS with the graphical method for the Theta angle in the supine and orthostatic positions. We observed that there is a bimodal distribution of Theta angle (θ*s*: 51.09±26.43, Coefficient of variation: 0.51747) in all patients populations during the supine period (Fig. 4C).

**Fig 4.**
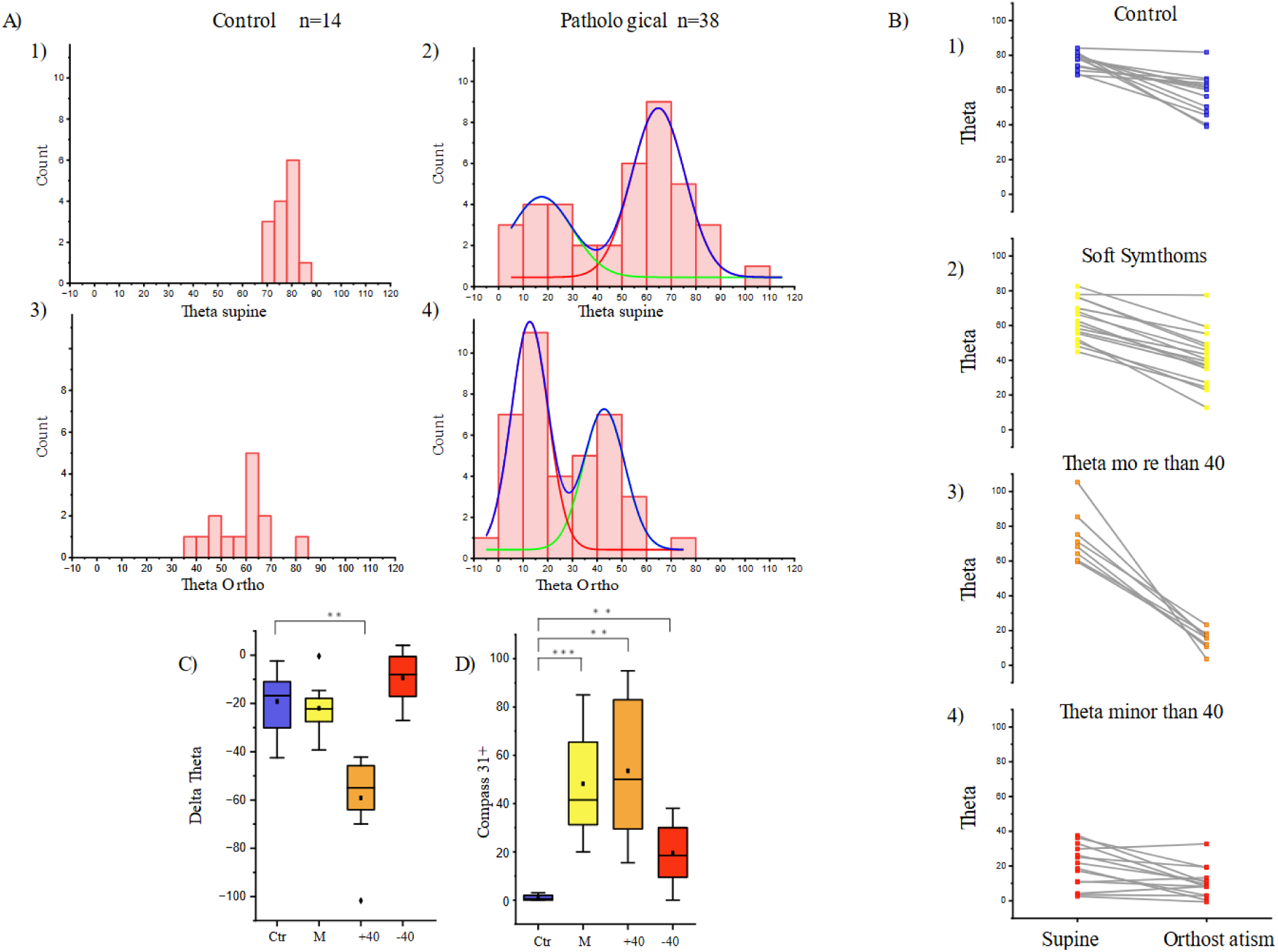
A) Histogram of the control and patient groups for the Theta value during supine and upright positions. A1) Histogram of the supine Theta measurements for the control group; the scale was adjusted to match that of the patient group. A2)Histogram of the supine Theta measurements for all patients; two Gaussian curves were adjusted above the graph, since it was observed that the data showed bimodality. A3)Histogram of the upright Theta measurement for the control group; the scale was adjusted to match that of the patient group. A4) Histogram of the upright Theta measurements for all patients; two Gaussian curves were adjusted above the graph, since it was observed that the data showed bimodality. B) We have placed the Theta[θ] values for the supine (Left) and orthostatism (Right) periods of all study subjects; these values were connected by a line to their corresponding value, and each group was assigned a color (blue for controls, yellow for MP-S, orange for MP Δθ>40 and red for the MP θ<40 group). B1) Theta values for the supine and upright positions of the control group. B.2) Theta values for the supine and upright position of the MP-S group. B3) Theta values for the supine and upright position of the MP Δθ>40 group. B4) Theta values for the supine and upright position of the MP θ<40 group. C) Box plot for the ΔTheta[θ] value of all groups, where each group was assigned a color (blue for controls, yellow for MP-S, orange for MP Δθ>40 and red for the MP θ<40 group). D) Box plot for the Compass 31+ score of the patients in each of the selected groups. To indicate statistical significance with respect to the control group, an asterisk [*] was placed for a p<0.05, two asterisks [**] for p<0.001 and tree asterisks [***] for p<0.0001.

Therefore, we observed that the Theta supine (θ*s*) values seemed to fit a bimodal distribution, and that the two populations have an intersection at the 40.651° of Theta value. Subsequently, we decided to perform a subtraction between Theta ortho(θ*o*) and Theta supine; this is a delta Theta change (Δθ = θ*o* − θ*s*) (Fig. 4).

Thus, we determined that the pathological groups could be classified into 3 distinct groups based on this division of the two populations of θ*s* and the Δθ, into the following: 1) Mild POTS with a Δθ change equal or greater than 40° (MP Δθ≥40°) n=8; 2) A group of Mild POTS with a Theta supine value (θ*s*) that starts below 40° and remains so during the upright position (MP θ<40 = θ*s*≤40° & θ*o*≤40°) n=14; 3) A third Mild POTS with OI symptoms that does not meet the above criteria; this has the softest change in Δθ (MP-S) n=16 (Fig. 4).

During the supine period, control group do not show significant differences with POTS subgroups in Systolic Blood Pressure (SBP), but during orthostatism, SBP in control group (control: 104.85±16.30) was significantly different from MP-S (115.78±13.94; p=0.04831), MP Δθ>40(127.61±23.45; p= 0.01854), and MP θ<40(130.59±20.10; p=0.00719) groups (Fig 5A).

**Fig. 5.**
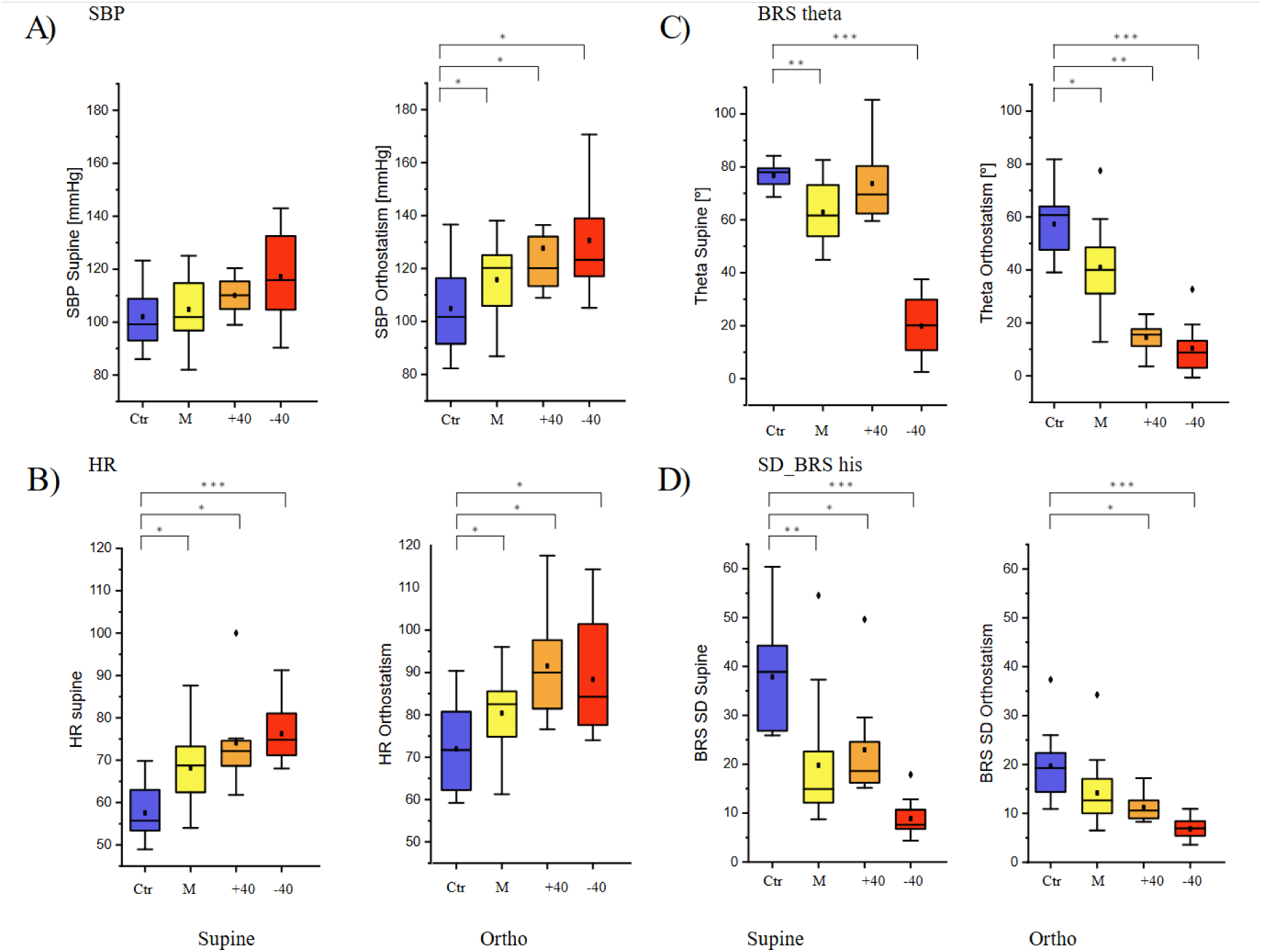
Box Plots for SBP(A), HR (B), Theta(C), and the SD of the BRS histogram (D) during the supine(left) and upright position(right). To indicate statistical significance with respect to the control group, an asterisk [*] was placed for a p<0.05, two asterisks [**] for p<0.001 and tree asterisks [***] for p<0.0001.

In HR control during the supine position, significant differences were observed (control: 57.56±6.72) compared to the MP-S (68.17±8.55; p=0.00736), MP Δθ>40 (74.08±11.27; p=0.00593) and MP θ<40 (76.25±6.55; p<0.001), as well as during orthostatism (control: 71.96±9.86; MP-S: 80.40±9.54, p= 0.01307; MP Δθ>40: 91.55±13.22, p= 0.00149; MP θ<40: 88.36±12.64, p= 0.00131) (Fig 5B).

The delta values in heart rate between supine and ortho (ΔHR) were not significantly different between healthy controls and POTS subgroups, while the ΔSBP for the MP-S (13.05±6.71; p= 0.00733) and MP Δθ>40 (11.11±12.72; p= 0.03156) groups were significantly different than those of control group patients (2.89±9.58) (Table 1).

### 3.3. BRS sensitivity decrease in LONG Covid patients

For the BRS measured by the Theta(θ) values during the supine period, the MP-S (62.90±11.53; p=0.00095) and MP θ<40 (19.81±12.11; p<0.0001) groups are significantly different than the control group(76.71±4.66), while the MP Δθ>40 group is not; however, during the orthostatic period, all groups mean values differed from the control group (57.31±11.68), MP-S (40.92±15.63; p=0.00296), MP Δθ>40 (14.48±5.85; p=0.00015) and MP θ<40 (10.37±8.84; p<0.0001) (Fig 5C).

#### 3.3.1. BRS dispersion measures

The mean of BRS histogram was lower in all patient groups in the supine period (control: 18.44±4.56; MP-S: 12.54±9.75, p=0.01502; MP Δθ>40: 7.15±4.88, p=0.00033; MP θ<40: 2.97±2.04, p<0.0001); in the upright position only the MP Δθ>40 (2.01±1.88; p=0.00056) and MP θ<40 (1.94±1.53; p<0.0001) patient groups had a significant difference compared to the healthy control group (7.43±3.41) (Table 1).

The standard deviation of BRS was also studied, since it was noticed in the individual’s histogram that patients had reduced variability and seemed to have a higher Kurtosis than those of the healthy individuals.

The standard deviation for all patient groups was significantly different from that of healthy individuals during the supine period (control: 37.87±10.54; MP-S: 19.77±12.16, p=0.00027; MP Δθ>40: 22.95±11.67, p=0.0086; MP θ<40: 8.87±3.56, p<0.0001), while in the upright position, only the standard deviation of the MP Δθ>40 (11.24±2.91; p=0.0015) and MP θ<40 (6.81±2.11; p<0.0001) groups were significantly different from healthy controls (19.67±6.85) (Fig. 3D).

### 3.4. POTS subgroups are supported by canonical HRV variables

In the supine position, all patient groups had significantly different values for the SDNN (Fig 6A left) (control: 65.60±18.16; MP-S: 39.07±18.85, p=0.00095; MP Δθ>40: 37.11±12.39, p=0.00118; MP θ<40: 20.28±7.90, p<0.001), RMSSD (Fig 6B left) (control: 77.35±20.33; MP-S: 45.85±25.26, p=0.00182; MP Δθ>40: 46.01±21.25, p=0.00858; MP θ<40: 16.88±5.13, p<0.0001) and pNN50 (Fig 6C left) (control: 47.93±12.41; MP-S: 24.07±24.01, p=0.00936; MP Δθ>40: 15.77±13.23, p=0.00563; MP θ<40: 0.81±1.61, p<0.0001). In addition, the MP θ<40 group had SDNN Values below 50 ms (Shaffer, 2017), RMSSD lower than 30 ms (Tegegne et al., 2019) and pNN50 lower than 3% (Corrales et al., 2012); all of which are considered high-risk values.

**Fig. 6.**
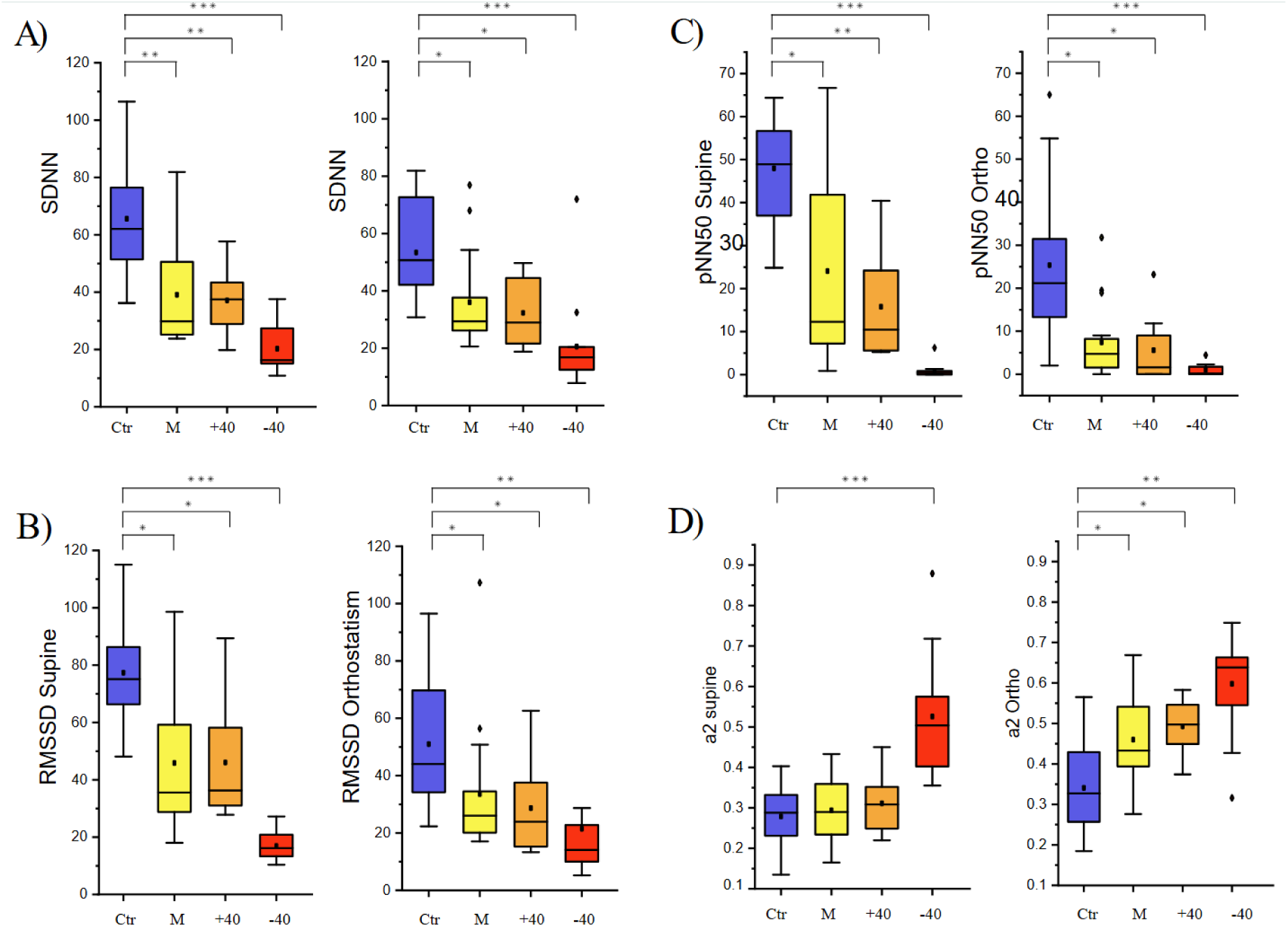
Box plots of the SDNN(A), RMSSD(B), pNN50(C) and **α**2(D) for each of the groups, in the supine(left) and upright(right) position. To indicate statistical significance with respect to the control group, an asterisk [*] was placed for a p<0.05, two asterisks [**] for p<0.001 and tree asterisks [***] for p<0.0001.

The values were also significantly different during the orthostatism period for the SDNN (Fig 6A right) (control: 53.36±17.18; MP-S: 36.02±16.34, p=0.00386; MP Δθ>40: 32.32±13.00, p=0.01537; MP θ<40: 20.54±15.95, p<0.0001), RMSSD (Fig 6B right) (control: 50.97±23.90; MP-S: 33.55±22.67, p=0.01502; MP Δθ>40: 28.66±16.88, p=0.02223; MP θ<40: 21.46±24.18, p=0.00309) and pNN50 (Fig 6C right) (control: 25.34±18.90; MP-S: 7.40±8.72, p=0.00338; MP Δθ>40: 5.5525±8.24167, p=0.00565; MP θ<40: 0.96±1.32, p<0.0001).

The Theta angle in the MP θ<40 group was defined by a low variability with a pNN50 range of 0-2.29[ms], a RMSSD range of 5.2-45.64[ms] and a collapsed ellipse area of 285.82-1400.79. The MP-S group had a lower pNN50 than those of the control; however, the MP θ<40 group was the one with the lowest values compared to the others.

### 3.5. Canonical variables correlate with new variables: HRV Variables vs BRS Theta

It is already known that the HRV variables decrease in POTS and Long-Covid patients (Hira et al., 2025, Teodorescu et al., 2025, Swai et al., 2019, Shah et al., 2022), but it is unknown how these variables are correlated with the BRS in the POTS groups. Therefore, we performed a scatter plot where the HRV variables were placed on the X axis and our proposed variables (Theta and BRS SD) were placed on the Y axis.

With the purpose of describing the data’s behaviour, we attempted to fit certain functions in order to explain the relation between both sets of variables.

Based on the observed pattern of the data, a S-shaped logistic function was fitted to supine graphs data, and since S-shaped did not fit properly to the orthostatic period graphs, we implemented a Logistic function.

The Theta value showed a strong correlation with all canonical variables; however, the strongest correlations appeared between the Theta value and the supine RMSSD (0.9 Spearman test) and pNN50 (0.85 Spearman test), suggesting that our Theta value has a significant influence on parasympathetic measurement. But also, the relationship between SDNN (0.83 Spearman test) and the Theta value is very strong, leading us to conclude that Theta can be associated with the individual’s autonomic response (Fig. 7).

**Fig 7.**
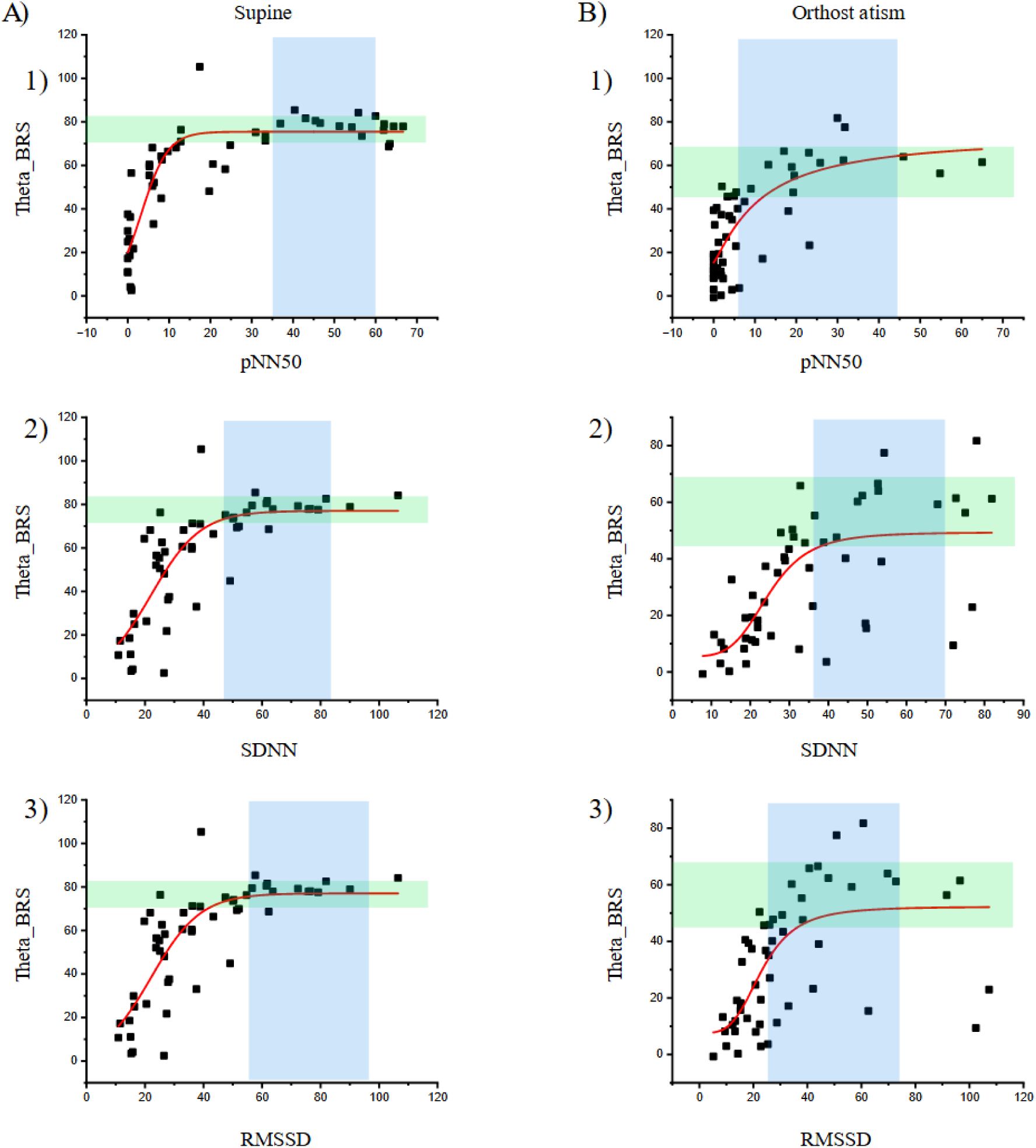
HRV variables have been placed on the X-axis to compare them with the Theta values, which have been placed on the Y-axis in a scatter plot. Additionally, the range of the control group has been highlighted with a blue and green rectangle. A1) Scatter plot of the pNN50 and Theta values during the supine position with an adjusted S-Logistic function. A2) Scatter plot of the SDNN and Theta values during the supine position with an adjusted S-Logistic function. A3) Scatter plot of the RMSSD and Theta values during the supine position with an adjusted S-Logistic function. B1) Scatter plot of the pNN50 and Theta values during the upright position with an adjusted Logistic function. B2) Scatter plot of the SDNN and Theta values during the upright position with an adjusted Logistic function. B3) Scatter plot of the RMSSD and Theta values during the upright position with an adjusted Logistic function.

#### HRV Variables vs SD BRS

The correlation between the SD BRS and the HRV variables is shown with a linear regression for SDNN during both positions, as well as for the RMSSD during the supine period.

For pNN50 during the supine position, a S-Logistic function was applied, and for the orthostatism period, a Logistic fit was performed. The Logistic fit was also applied to the RMSSD during the upright position (Fig. 8).

**Fig. 8.**
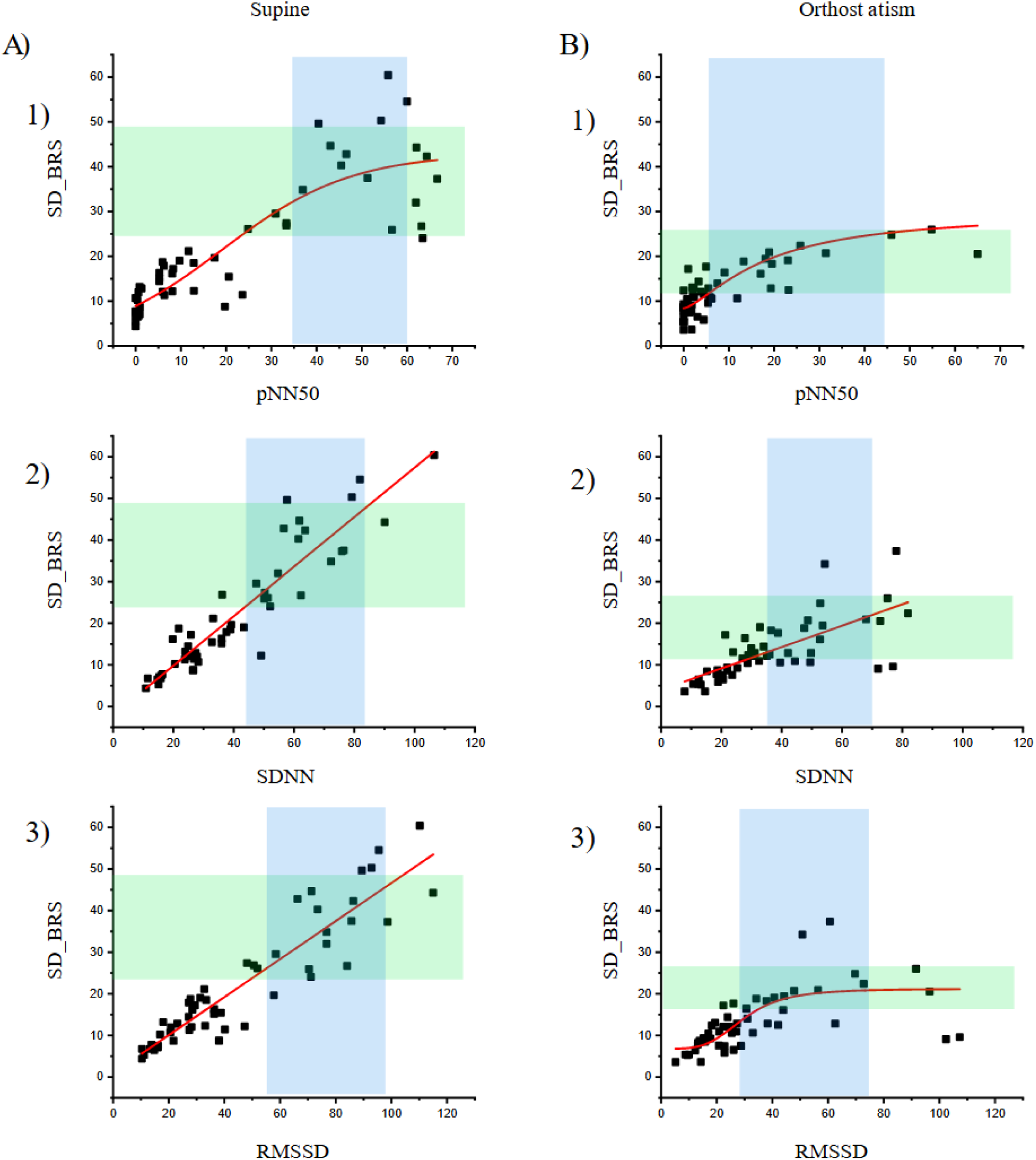
HRV variables have been placed on the X-axis to compare them with the SD BRS values obtained, which have been placed on the Y-axis in a scatter plot. Additionally, the range of the control group has been highlighted with a blue and green rectangle. A1) Scatter plot of the pNN50 and SD BRS values during the supine position with an adjusted S-Logistic function. A2) Scatter plot of the SDNN and SD BRS values during the supine position with an adjusted linear function. A3) Scatter plot of the RMSSD and SD BRS values during the supine position with an adjusted linear function. B1) Scatter plot of the pNN50 and SD BRS values during the upright position with an adjusted Logistic function. B2) Scatter plot of the SDNN and SD BRS values during the upright position with an adjusted linear function. B3) Scatter plot of the RMSSD and SD BRS values during the upright position with an adjusted Logistic function.

To determine how the patient groups correlate with the HRV values, we decided to make graphs with 3 axes corresponding to our variables and the HRV variables, in order to describe this progression of BRS dysfunction (Fig. 9).

**Fig. 9.**
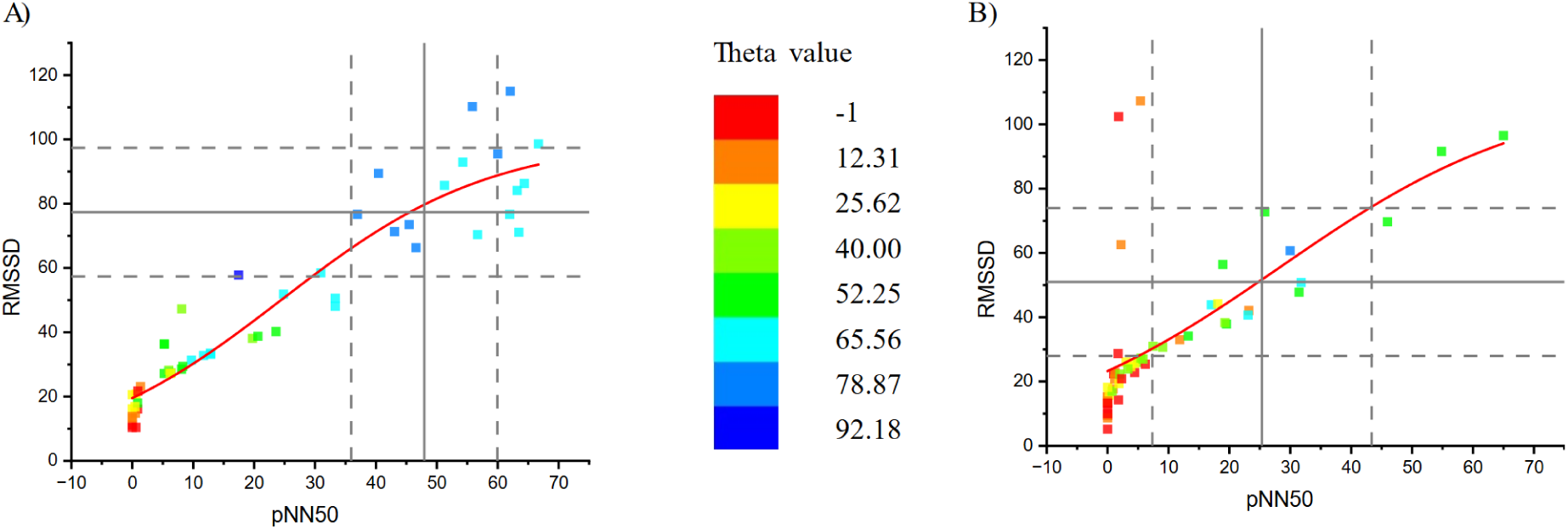
A) Scatter plot where pNN50 supine for all groups has been placed over the X-axis and the corresponding RMSSD supine value has been placed in the Y-axis.B) Scatter plot where pNN50 for orthostatism of all groups has been placed over the X-axis and the corresponding RMSSD for orthostatism value has been placed in the Y-axis. A single color gradient has been applied on both graphs.

It is important to note that the intersection of the two standard deviations of the controls on the two axis ended up close to the Theta value of 40, as we mentioned above (green color); this value divides the subgroups.

Furthermore, a Spearman correlation test was performed for the new proposed variables and the canonical HRV variables; this resulted in a very strong correlation, where most of them had a positive correlation of over 0.5. We conducted a Spearman test since we observed that the correlations did not have a linear behaviour with each other (Fig. 10).

**Fig. 10.**
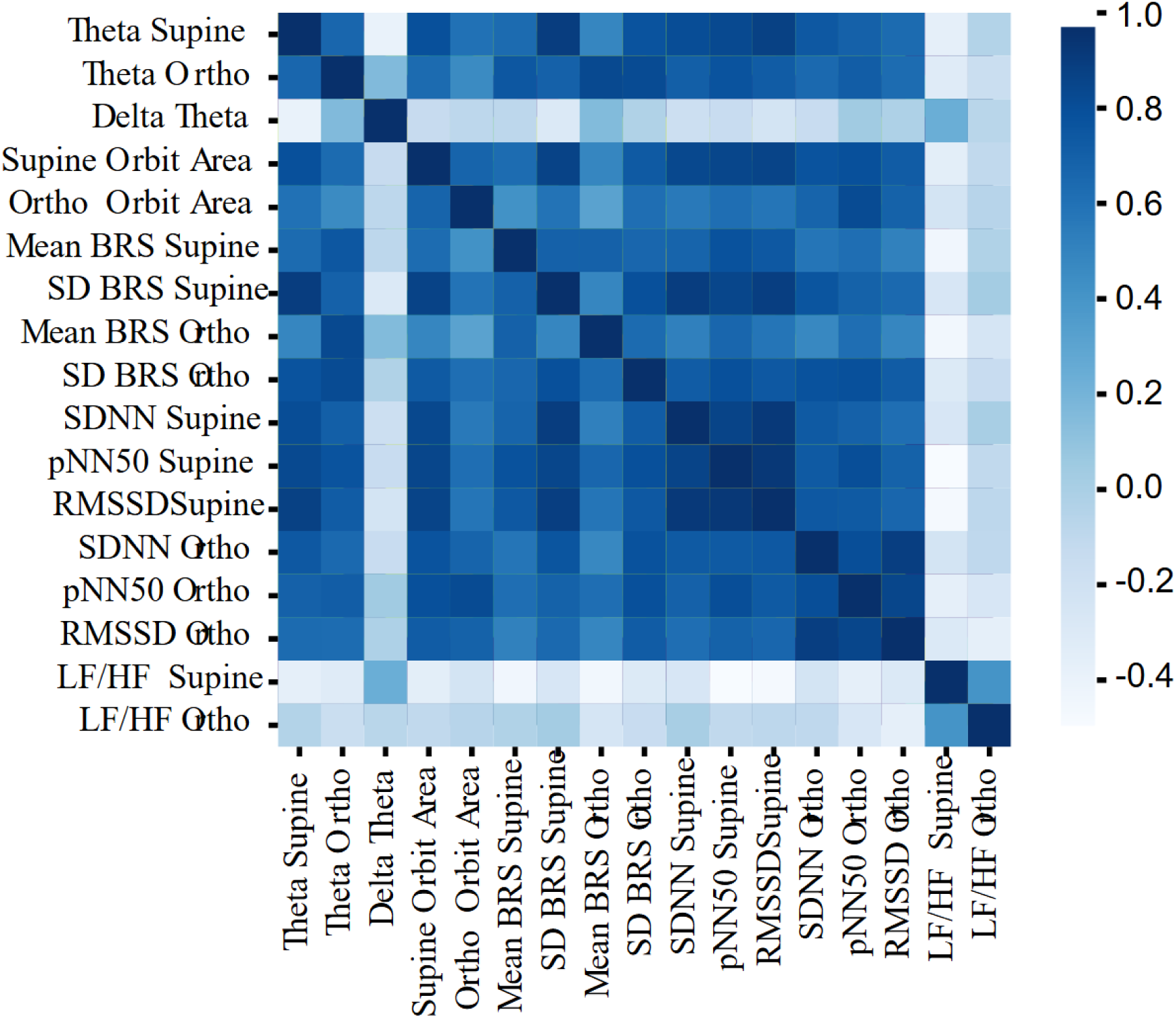
Matrix with the canonical HRV and proposed Theta and SD BRS values during the supine and upright positions, where a heatmap was applied to match the correlation values obtained using the Spearman test.

To summarize, when looking at the percentage change in BRS compared to the control group, we observed a decrease of 17.99% for the MP-S, 3.97% for MP Δθ>40, and 74.17% in the MP θ<40 group for the supine position. During orthostatism, the percentage drop for the control group is 25.28%, while for the patient groups it is 46.65% for the MP-S, 81.11% for MP Δθ>40, and 86.47% in the MP θ<40 group.

In conclusion, there is a drop in the BRS function of all groups; however, the MP Δθ>40 has the highest change and these are the patients with the most and worst symptoms. On the other hand, MP θ<40 patients have less symptoms, but these are more severe.

## 4. Discussion

Although none of our patients met the criteria for Hyperadrenergic POTS previously described, they all continued to present with symptoms ranging from minor to severe. Although no direct correlation was found between increased SBP or HR and symptom severity, this prompted us to look for other variables that might explain our patients’ mild POTS symptoms.

It has been previously reported that patients who suffer from OI tend to have depressed/attenuated cardiac vagal baroreflex sensitivity (Farquhar et al., 2000; Mustafa et al., 2012; Stewart et al., 2021), and our data show different degrees of BRS decrease among all Long covid patients. This deficiency is strongly supported by the dual decrement of the Theta and SD BRS values.

In healthy individuals, there is a proportional change in the IBI measurement for every 1 unit of SBP change, this is referred to as a change of ∽5-10 ms per mmHg (de Boer & Karemaker, 2019; Svčinová et al., 2015). However, diabetic patients have previously shown a decrease in baroreflex sensitivity of about 30-50% (Frattola et al., 1997; Lefrandt et al., 1999; Michel-Chávez et al., 2015), due to an autonomic dysfunction caused by structural damage to the vagal afferent and the brainstem control; this even before a fully diagnosed diabetic state (Svčinová et al., 2013, Cseh et al., 2020). These patients are known to have a mild BRS dysfunction (Rowaiye et al., 2013).

Similar to the mild BRS dysfunction present in diabetic patients, our mild POTS patients have also shown different degrees of BRS decrement.

On the other hand, viral infections such as HPV damage the brainstem functions, and also elicit POTS-like symptoms (Brinth et al., 2015; Blitshteyn et al., 2014b).

Likewise, SARS-CoV-2 infections damage the brainstem and cause a neuroinflammatory response (Matshchke et al., 2020; Emmi et al., 2023). The neuroinflammation could affect the firing rate of local brainstem nuclei and the baroreceptor adaptation; therefore, this impairs the correction factor (error signal), which could be the cause for the POTS-like symptoms (Blitshteyn et al., 2025a; Shi et al., 2011; Zubcevic et al., 2013).

In our patients, the BRS drop is around 80% in the group with the strongest symptoms; this means less adaptability to postural changes, leading to an impaired cardiac vagal baroreflex and autonomic nervous system imbalance.

Here we present a new graphical method that can be used to estimate BRS and that does not necessarily require an algorithm or software to be evaluated; in addition, it directly generates a graph that can be interpreted immediately through the Theta angle [θ] and the area of the ellipse. As previously demonstrated by Spearman’s correlation, these new variables also give information that correlates with changes in the HRV.

The analysis that has been described allows us to recognize as pathological those patients who have symptoms but do not meet the diagnostic criteria; this means that they can receive treatment that better aligns with their pathological group.

As we have shown before, those in the MP-S group still have an overall active cardiac vagal Baroreflex, which means that this group has the highest probability of responding positively to a possible treatment, by reversing the symptoms and effects of the disease.

Meanwhile, the MP Δθ>40 and MP θ<40 groups exhibit a BRS attenuation and a reduced ellipse area. Especially, the MP θ<40 is the group that shows the greatest area reduction, which can be interpreted as the closest group to baroreflex failure.

In conclusion, the subgroups presented show how the BRS impairment could be progressive, and its relationship with symptoms and alterations in HRV; this could be important for developing further treatment and diagnostic methods.

## Data Availability

All data produced in the present study are available upon reasonable request to the authors.

