## Supplementary material for "A new method to measure Baroreflex sensitivity impairment in Long Covid patients with Hyperadrenergic POTS-like symptoms": Table 1

| Variable | Control | | MP-S | | MP Δθ>40 | | MP θ<40 | | p Controls | | | p groups | | |
|---|---|---|---|---|---|---|---|---|---|---|---|---|---|---|
| | Mean | SD | Mean | SD | Mean | SD | Mean | SD | MP-S | MP Δθ>40 | MP θ<40 | MP-S - MP Δθ>40 | MP-S - MP θ<40 | MP Δθ>40 - MP θ<40 |
| SBP Sup | 102.01857 | 11.1525 | 104.78842 | 13.21291 | 110.03032 | 7.4193 | 117.15441 | 16.97713 | 0.49874 | 0.06053 | 0.05654 | 0.41974 | 0.10171 | 0.60873 |
| SBP ort | 104.85816 | 16.30694 | 115.78017 | 13.94011 | 127.61636 | 23.45853 | 130.59447 | 20.10061 | 0.04831* | 0.01854* | 0.00719* | 0.52023 | 0.23611 | 0.8112 |
| D SYS | 2.89502 | 9.58423 | 13.05299 | 6.71733 | 18.77822 | 21.0162 | 11.11702 | 12.72665 | 0.00733* | 0.03156* | 0.11292 | 0.87832 | 0.28912 | 0.56182 |
| HR sup | 57.56538 | 6.7291 | 68.17216 | 8.55282 | 74.08051 | 11.27957 | 76.25174 | 6.55669 | 0.00147* | 0.00119* | <0.0001* | 0.20935 | 0.00647* | 0.20671 |
| HR ort | 71.9631 | 9.86144 | 80.40115 | 9.5452 | 91.55044 | 13.22251 | 88.36685 | 12.6461 | 0.02615* | 0.00299* | 0.00262* | 0.07084 | 0.20483 | 0.56182 |
| D Hr | 14.05016 | 5.34415 | 12.25368 | 4.97418 | 17.87958 | 5.85773 | 12.19448 | 8.09406 | 0.30845 | 0.10873 | 0.24133 | 0.02971* | 0.60332 | 0.10873 |
| Escent. IBI su | 0.6524 | 0.09432 | 0.6085 | 0.17156 | 0.5794 | 0.23483 | 0.82859 | 0.05607 | 0.51935 | 0.8112 | <0.0001* | 0.92681 | <0.0001* | 0.000439759* |
| Escent. IBI ort | 0.80627 | 0.07532 | 0.79878 | 0.08284 | 0.84992 | 0.03484 | 0.8316 | 0.05883 | 0.88427 | 0.14203 | 0.26994 | 0.13353 | 0.23611 | 0.51673 |
| Sd2/Sd1 SUP | 1.83115 | 0.35655 | 1.85894 | 0.64131 | 1.71132 | 0.44729 | 3.64826 | 1.03102 | 0.60332 | 0.60873 | <0.0001* | 0.73626 | <0.0001* | 0.000199417* |
| Sd2/Sd1 ORT | 3.12613 | 0.84913 | 3.22175 | 1.20858 | 3.58568 | 0.6673 | 3.48114 | 0.99138 | 0.98342 | 0.2601 | 0.44837 | 0.28388 | 0.41758 | 0.75874 |
| Theta Sup | 76.71298 | 4.663 | 62.90532 | 11.53582 | 73.66016 | 15.32481 | 19.81135 | 12.11007 | 0.000950243* | 0.16176 | <0.0001* | 0.11839 | <0.0001* | 0.000151861* |
| Theta Ort | 57.31859 | 11.68058 | 40.92423 | 15.63466 | 14.4857 | 5.85119 | 10.37529 | 8.84247 | 0.00296* | 0.000151861* | <0.0001* | 0.000429693* | <0.0001* | 0.10873 |
| D Theta | -19.21487 | 12.52344 | -21.9811 | 8.48014 | -59.17446 | 19.37918 | -9.43606 | 10.45116 | 0.39411 | 0.000199417* | 0.05654 | 0.000100839* | 0.00195* | 0.000151861* |
| Theta Area su | 3485.47429 | 1384.14066 | 1663.35444 | 1236.926 | 1998.45763 | 2005.41555 | 760.43186 | 413.30994 | 0.000950243* | 0.01854* | <0.0001* | 0.87832 | 0.00296* | 0.01539* |
| Theta Area ort | 2224.996 | 1109.87403 | 1211.59238 | 504.56571 | 1738.39963 | 1281.81164 | 862.37321 | 538.42046 | 0.0057* | 0.2601 | 0.000439758* | 0.78288 | 0.05856 | 0.05175 |
| BRS Mean sup | 18.44325 | 4.56965 | 12.54794 | 9.75712 | 7.15853 | 4.88615 | 2.97578 | 2.04127 | 0.01502* | 0.000339347* | <0.0001* | 0.11839 | 0.000120417* | 0.01539* |
| BRS Mean ort | 7.43931 | 3.41137 | 5.79554 | 2.67385 | 2.01018 | 1.8806 | 1.94611 | 1.53069 | 0.12914 | 0.000567385* | <0.0001* | 0.000675324* | <0.0001* | 0.43251 |
| BRS SD Sup | 37.87677 | 10.54693 | 19.77632 | 12.16024 | 22.9589 | 11.67461 | 8.87212 | 3.56681 | 0.000275379* | 0.0086* | <0.0001* | 0.20935 | 0.000168469* | 0.000339347* |
| BRS SD ort | 19.67343 | 6.85517 | 14.18314 | 6.64112 | 11.24071 | 2.91722 | 6.81357 | 2.11188 | 0.01338 | 0.0015* | <0.0001* | 0.25726 | <0.0001* | 0.0019 |
| BRS Kurt sup | 5.44745 | 4.87857 | 3.40679 | 2.82398 | 3.85726 | 3.53722 | 6.88812 | 11.57615 | 0.15152 | 0.2601 | 0.73039 | 1 | 0.23611 | 0.20671 |
| BRS Kurt ort | 4.48993 | 3.71803 | 5.00736 | 5.75414 | 5.81131 | 5.66142 | 9.82521 | 19.29971 | 0.91723 | 0.91846 | 0.73039 | 1 | 0.60332 | 0.70737 |
| LF/HF Supine | 0.8915 | 0.53593 | 1.58963 | 3.89207 | 0.99025 | 1.06091 | 3.34221 | 2.70575 | 0.27057 | 0.70729 | 0.000864703* | 0.4082 | 0.000948997* | 0.00569* |
| LF/HF ORT | 2.03164 | 1.42173 | 1.73238 | 1.55982 | 2.13425 | 1.94884 | 2.59464 | 2.07395 | 0.46693 | 1 | 0.56573 | 0.64603 | 0.37145 | 0.75874 |
| SDNN SUP | 65.60714 | 18.16338 | 39.075 | 18.85352 | 37.1125 | 12.39798 | 20.28571 | 7.90442 | 0.000950243* | 0.00118* | <0.0001* | 0.97557 | 0.00296* | 0.00371* |
| SDNN ORT | 53.36429 | 17.18392 | 36.025 | 16.3413 | 32.325 | 13.00382 | 20.54286 | 15.9596 | 0.00386* | 0.01537* | <0.0001* | 0.52014 | 0.000322834* | 0.00568* |
| RMSSD SUP | 77.35714 | 20.33359 | 45.85625 | 25.26038 | 46.0125 | 21.25011 | 16.88571 | 5.13448 | 0.00182* | 0.00858* | <0.0001* | 0.78283 | <0.0001* | 0.000150558* |
| RMSSD ORT | 50.97143 | 23.90402 | 33.55625 | 22.67027 | 28.6625 | 16.88955 | 21.46429 | 24.18164 | 0.01502* | 0.02223* | 0.000309297* | 0.44399 | 0.00295* | 0.07042 |
| pNN50 SUP | 47.93857 | 12.41711 | 24.07125 | 24.01147 | 15.775 | 13.23706 | 0.81929 | 1.61303 | 0.00936* | 0.000563308* | <0.0001* | 0.4439 | <0.0001* | 0.000293153* |
| pNN50 ORT | 25.34571 | 18.90306 | 7.40438 | 8.72382 | 5.5525 | 8.24167 | 0.96714 | 1.32793 | 0.00338* | 0.00565* | <0.0001* | 0.35728 | 0.00091579* | 0.25148 |
