## Supplementary material for "A new method to measure Baroreflex sensitivity impairment in Long Covid patients with Hyperadrenergic POTS-like symptoms": Figures and captions

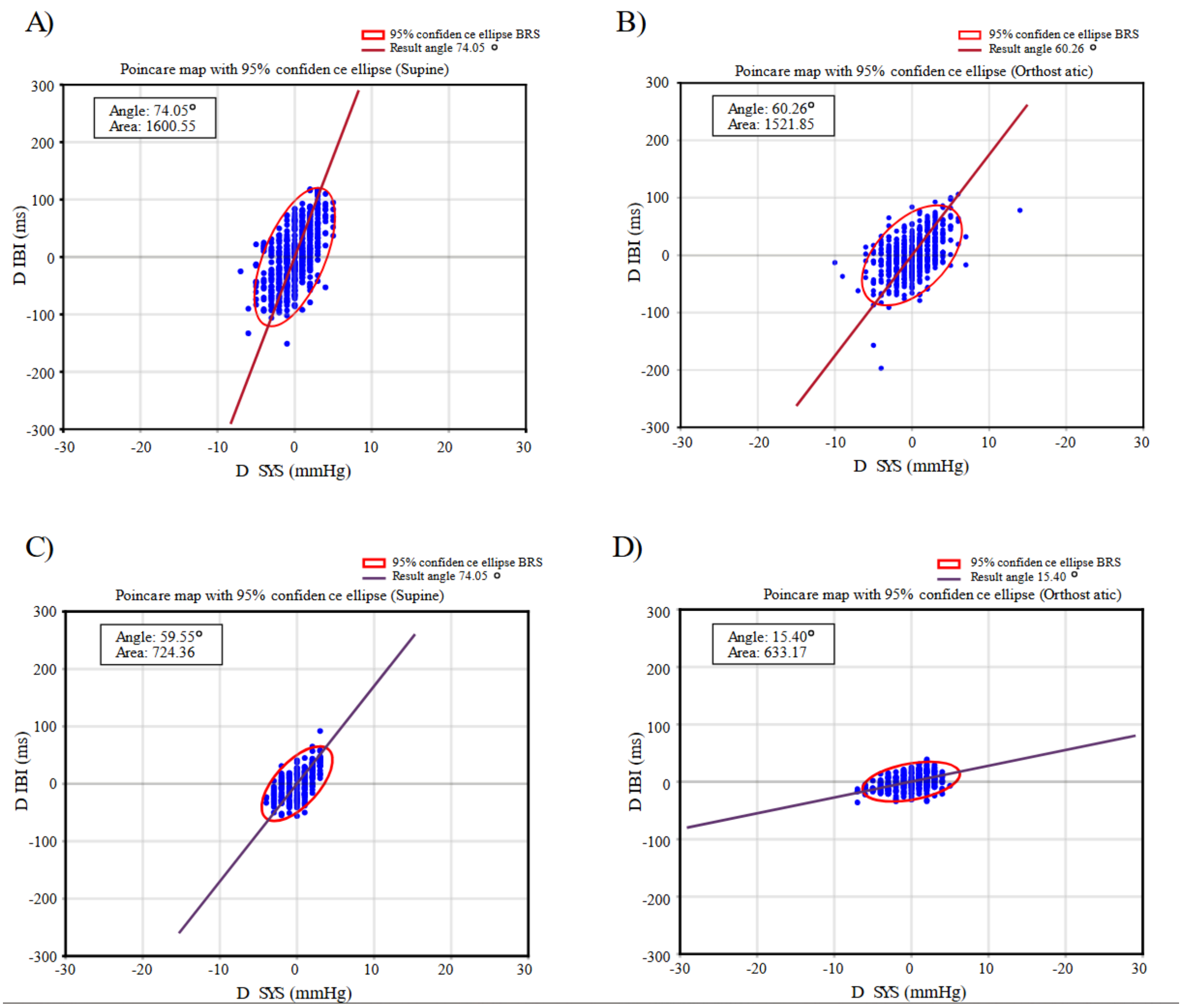


Fig1

The Theta graphics corresponding to the supine(A) and orthostatic(B) positions of a healthy subject is shown. We show the scatter plot of the ΔSBP and ΔIBI values in blue, over it a 95% confidence ellipse was adjusted and is shown in red; additionally, a dark red line with the calculated Theta[θ] angle was overimposed. The axis scales were normalized to observe the natural response without modifying the raw delta(Δ) values. 1A) Theta graph during the supine period for a control subject. 1B) Theta graph during the orthostatism period for a control subject. 1C) Theta graph during the supine period for a pathological subject. 1D) Theta graph during the orthostatism period for a pathological subject.

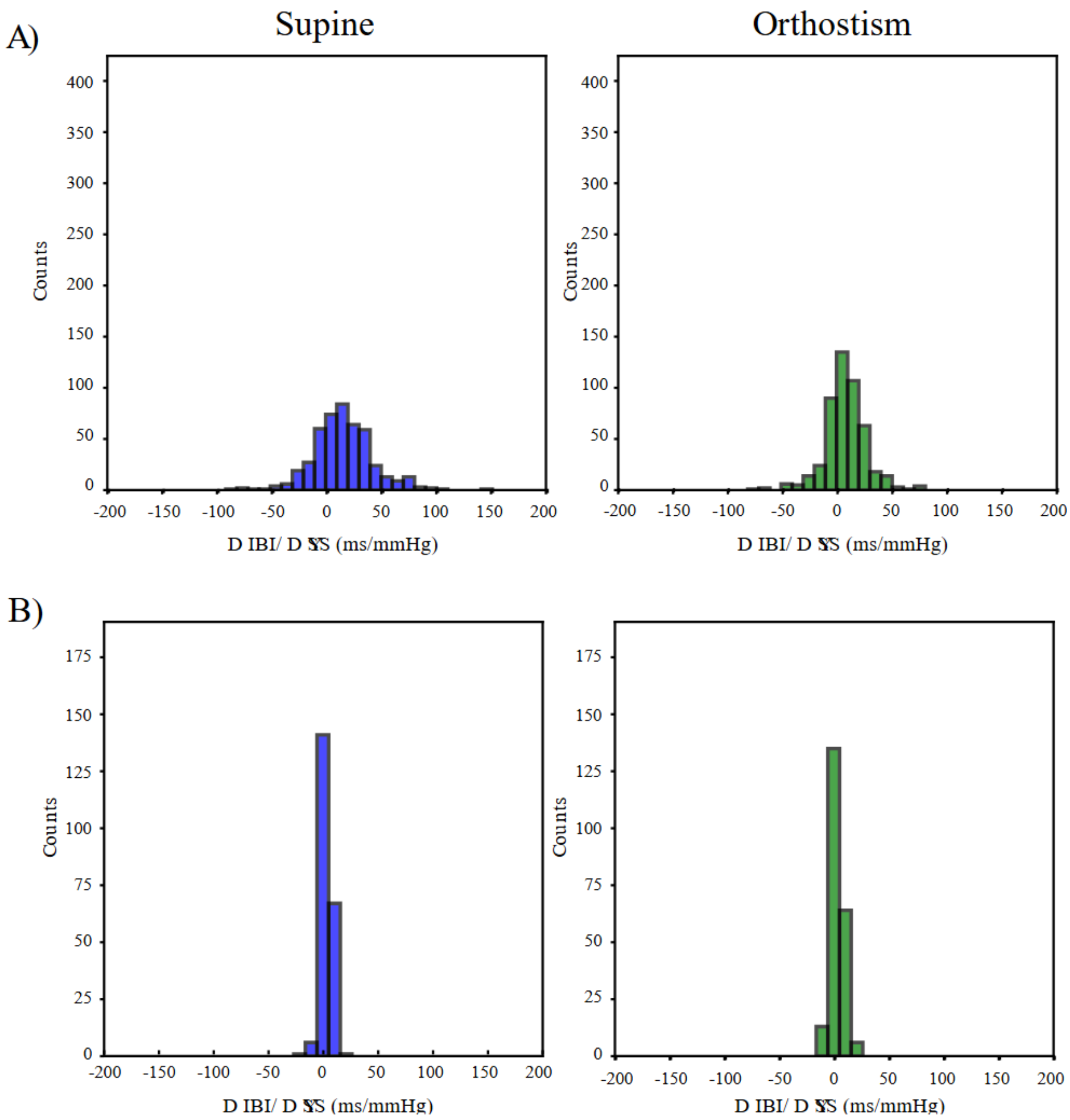


Fig2
Histogram for the BRS values corresponding to the supine (left) and orthostatic (right) positions, obtained from the division of ΔIBI/ΔSYS beat-to-beat measurements. 2A) Histogram of the BRS values for a control subject during the supine and upright positions. 2B) Histogram of the BRS values for a pathological subject during the supine and upright positions.

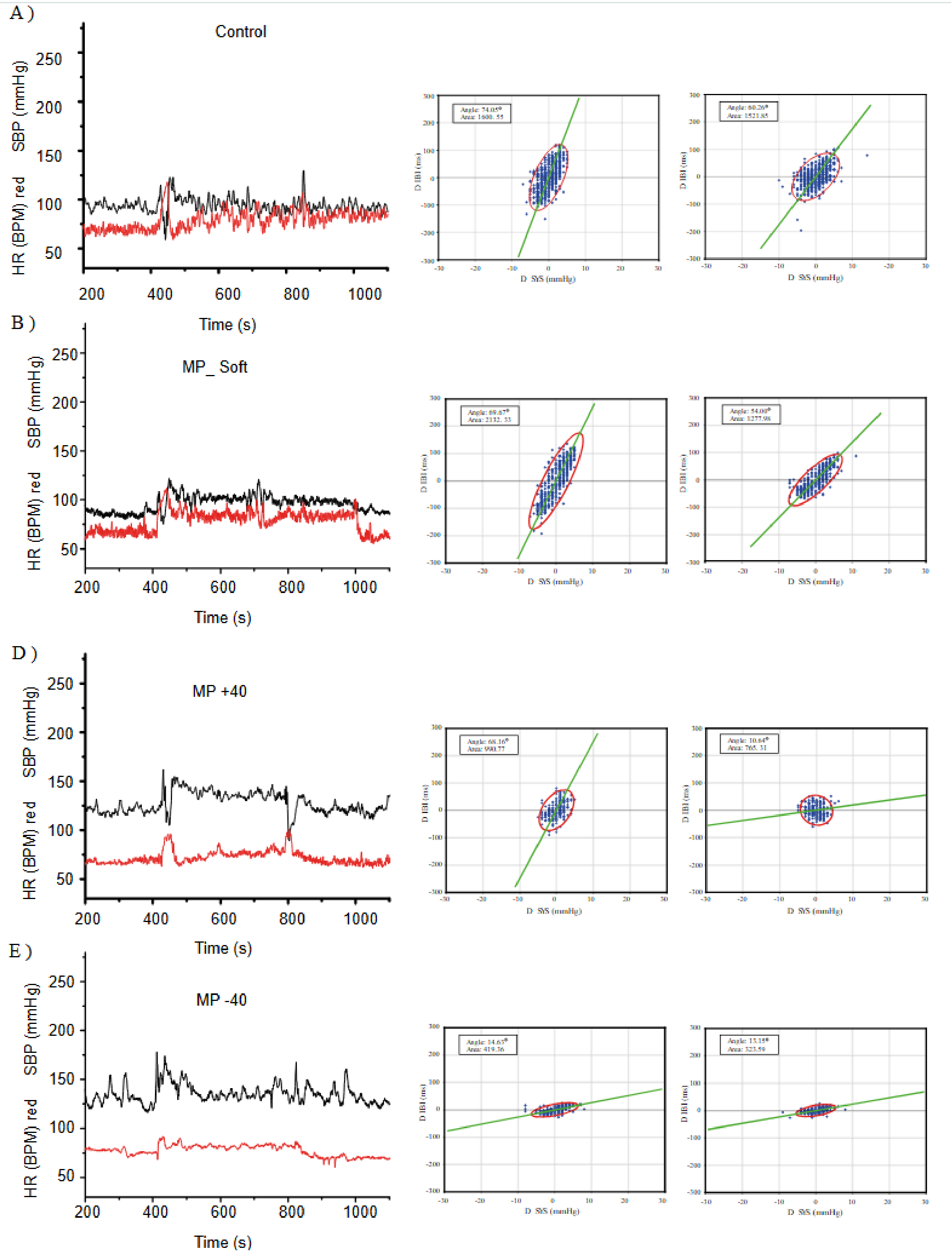


Fig3
Measured SBP and HR values (colored in black and red, respectively) of one patient from each of our selected groups, along with the BRS response represented by the Theta angle for the supine and upright positions. 3A) SBP and HR values of a healthy control accompanied by their respective Theta graphs. 3B) SBP and HR values accompanied by their respective Theta graphs of a patient with Mild POTS

and a soft change in the BRS response. 3C) SBP and HR values accompanied by their respective Theta graphs of a patient with Mild POTS and a change in angle >40º for the BRS response. 3D) SBP and HR values accompanied by their respective Theta graphs of a patient with Mild POTS and an attenuated BRS response.

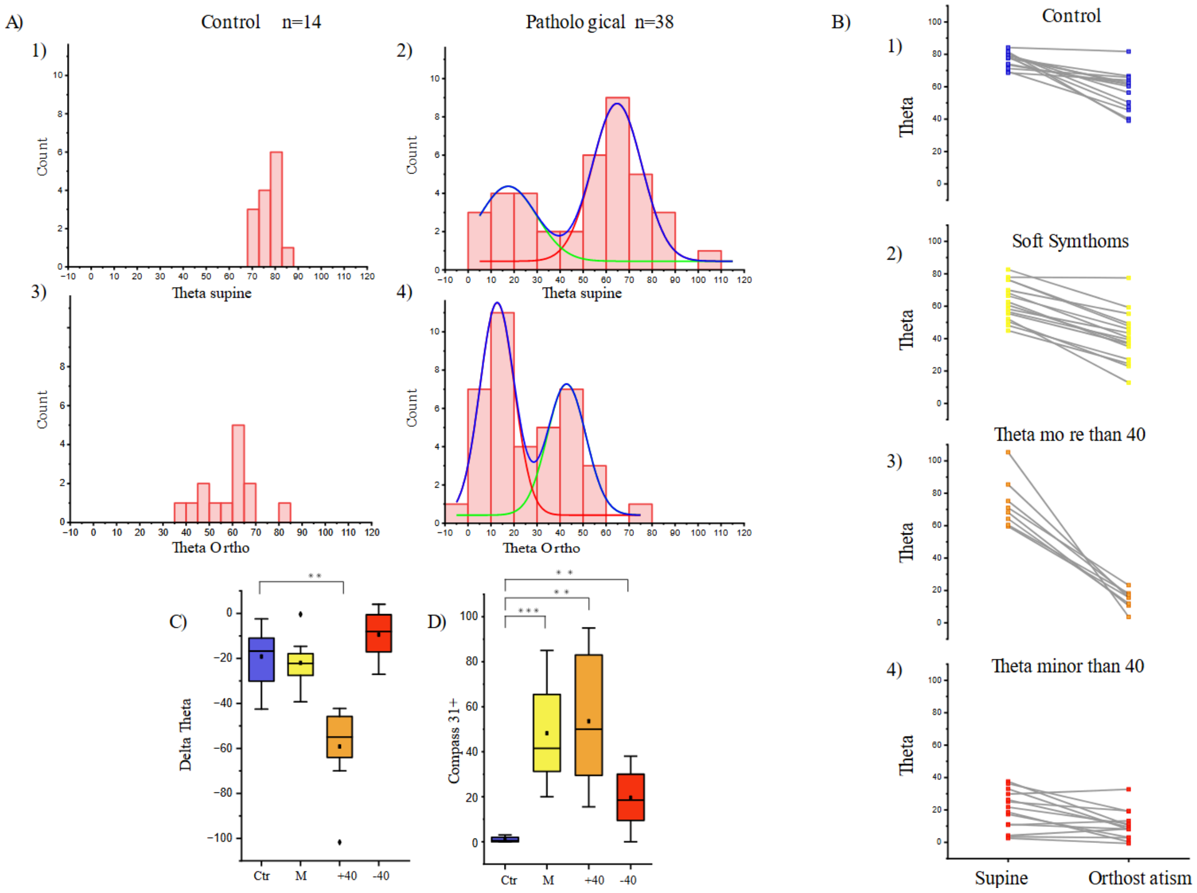


Fig 4

A) Histogram of the control and patient groups for the Theta value during supine and upright positions. A1) Histogram of the supine Theta measurements for the control group; the scale was adjusted to match that of the patient group. A2)Histogram of the supine Theta measurements for all patients; two Gaussian curves were adjusted above the graph, since it was observed that the data showed bimodality. A3)Histogram of the upright Theta measurement for the control group; the scale was adjusted to match that of the patient group. A4) Histogram of the upright Theta measurements for all patients; two Gaussian curves were adjusted above the graph, since it was observed that the data showed bimodality. B) We have placed the Theta[θ] values for the supine (Left) and orthostatism (Right) periods of all study subjects; these values were connected by a line to their corresponding value, and each group was assigned a color (blue for controls, yellow for MP-S, orange for MP Δθ>40 and red for the MP θ<40 group). B1) Theta values for the supine and upright positions of the control group. B.2) Theta values for the supine and upright position of the MP-S group. B3) Theta values for the supine and upright position of the MP Δθ>40 group. B4) Theta values for the supine and upright position of the MP θ<40 group. C) Box plot for the ΔTheta[θ] value of all groups, where each group was assigned a color (blue for controls, yellow for MP-S, orange for MP Δθ>40 and red for the MP θ<40 group). D) Box plot for the Compass 31+ score of the patients in each of the selected groups. To the indicate statistical significance with respect to the control group, an asterisk [*] was placed for a p<0.05, two asterisks [**] for p<0.001 and tree asterisks [***] for p<0.0001.

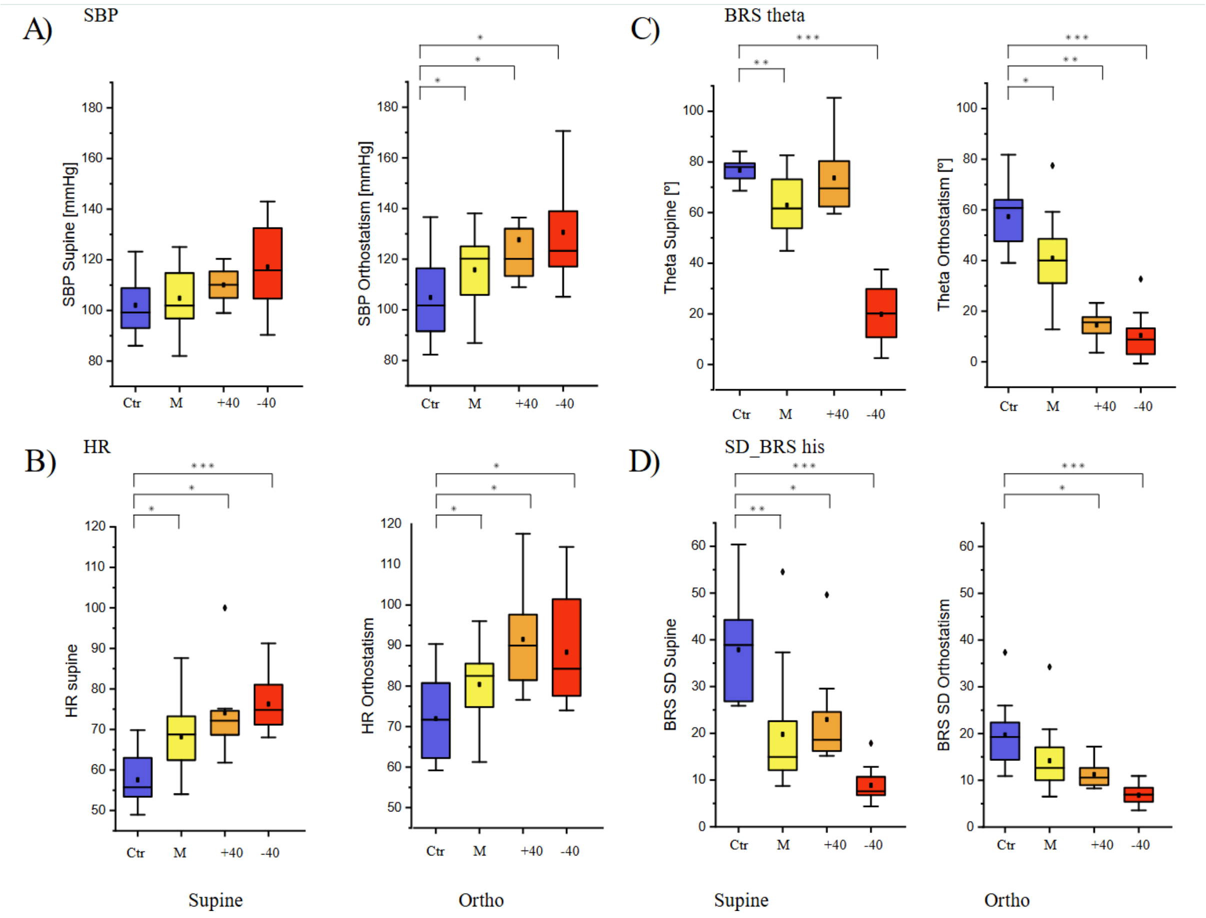


Fig 5

Box Plots for SBP(A), HR (B), Theta(C), and the SD of the BRS histogram (D) during the supine(left) and upright position(right). To the indicate statistical significance with respect to the control group, an asterisk [*] was placed for a p<0.05, two asterisks [**] for p<0.001 and tree asterisks [***] for p<0.0001.

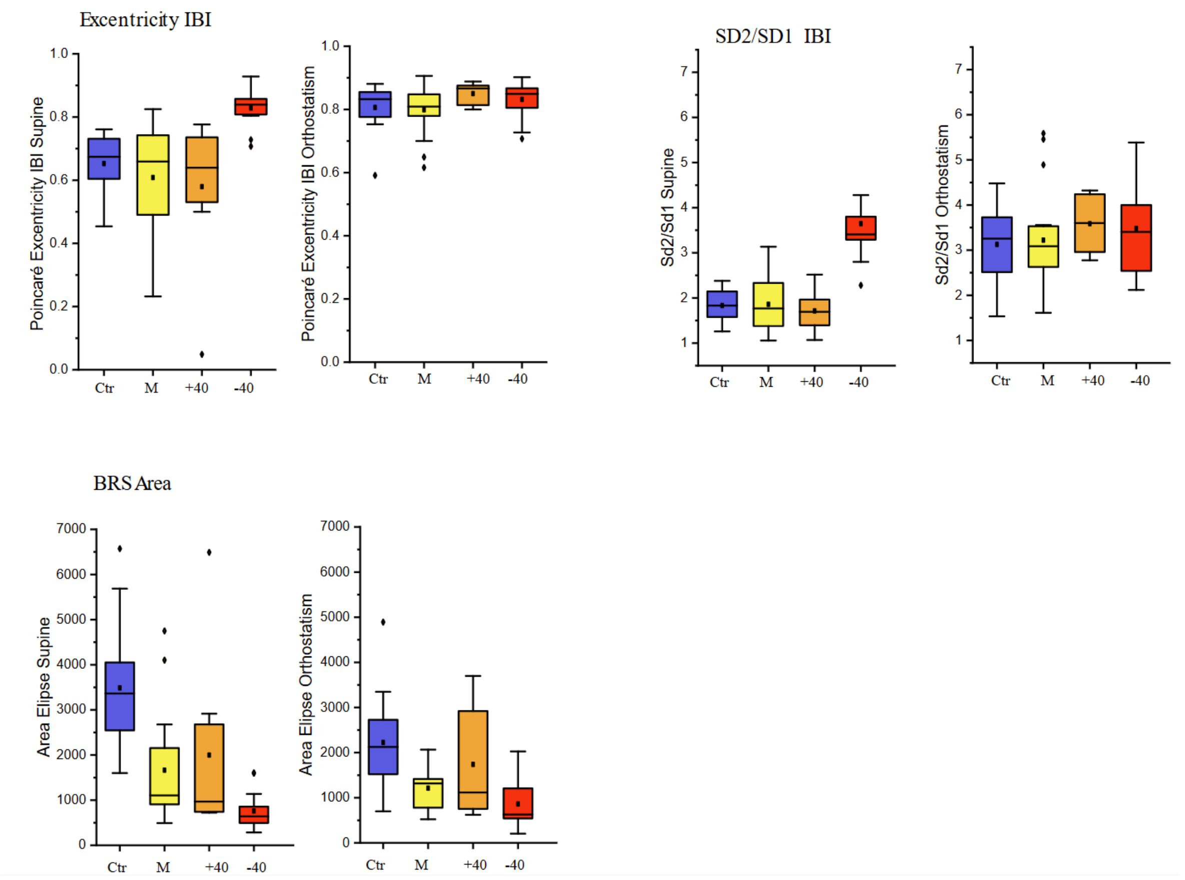


Supplementary Fig 1

Box plots with the data from Poincaré plots of each patient, such as eccentricity(A) and Sd2/Sd1(B), additionally the plot for the Area of the Theta ellipse(C) is also shown. To the indicate statistical significance with respect to the control group, an asterisk [*] was placed for a p<0.05, two asterisks [**] for p<0.001 and tree asterisks [***] for p<0.0001.

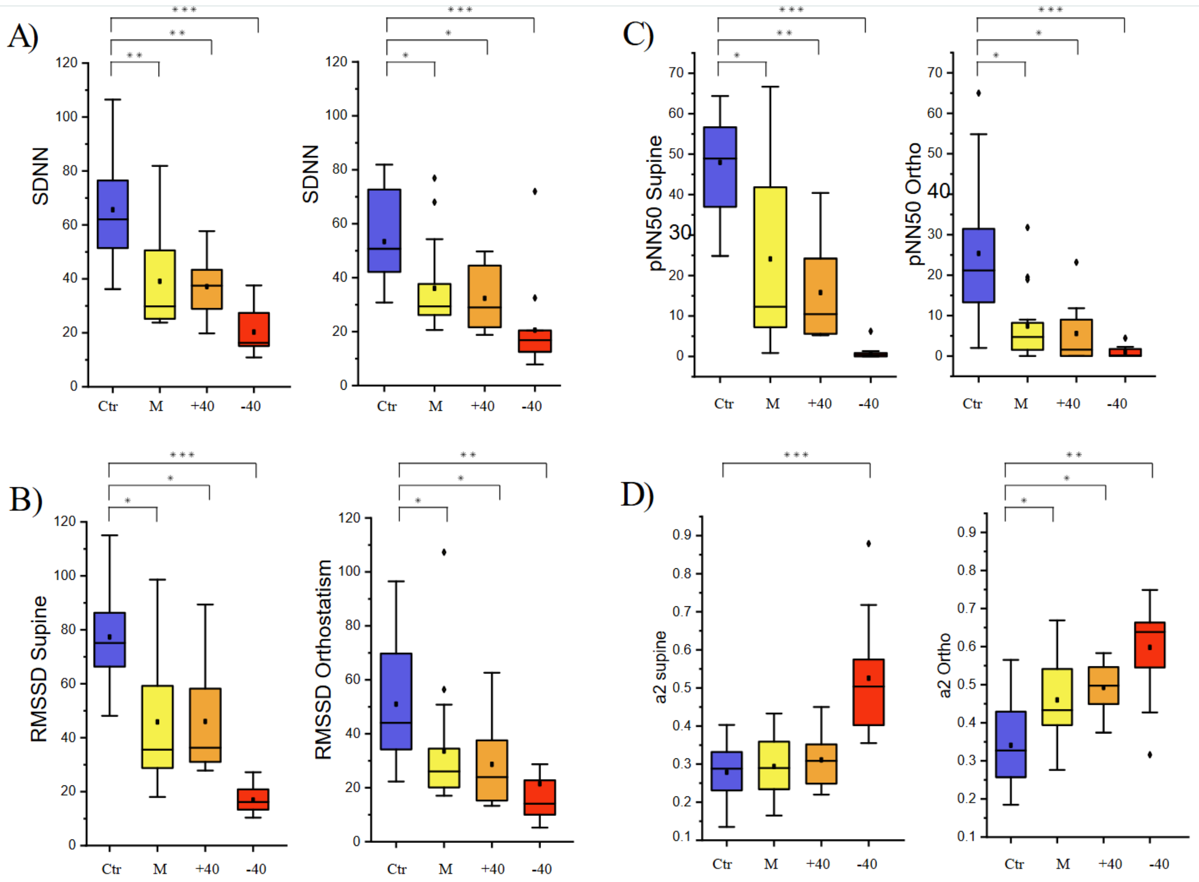


Fig 6.

Box plots of the SDNN(A), RMSSD(B), pNN50(C) and $\alpha 2$(D) for each of the groups, in the supine(left) and upright(right) position. To the indicate statistical significance with respect to the control group, an asterisk [*] was placed for a $p<0.05$, two asterisks [**] for $p<0.001$ and tree asterisks [***] for $p<0.0001$.

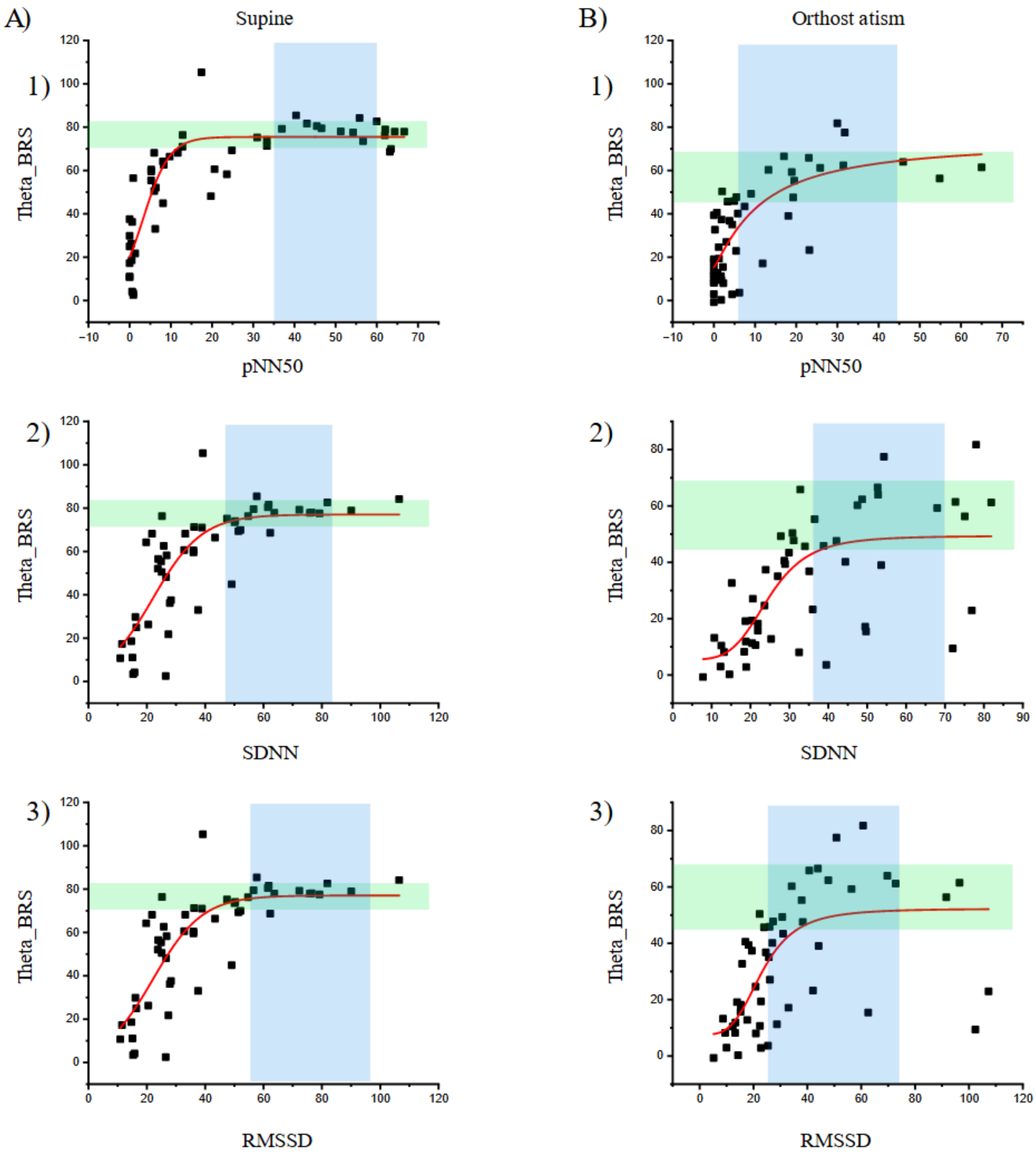


Fig 7
HRV variables have been placed on the X-axis to compare them with the Theta values, which have been placed on the Y-axis in a scatter plot. Additionally, the range of the control group has been highlighted with a blue and green rectangle. A1) Scatter plot of the pNN50 and Theta values during the supine position with an adjusted S-Logistic function. A2) Scatter plot of the SDNN and Theta values during the supine position with an adjusted S-Logistic function. A3) Scatter plot of the RMSSD and Theta values during the supine position with an adjusted S-Logistic function. B1) Scatter plot of the pNN50 and Theta values during the upright position with an adjusted Logistic function. B2) Scatter plot of the SDNN and Theta values during the upright position with an adjusted Logistic function. B3) Scatter plot of the

RMSSD and Theta values during the upright position with an adjusted Logistic function.

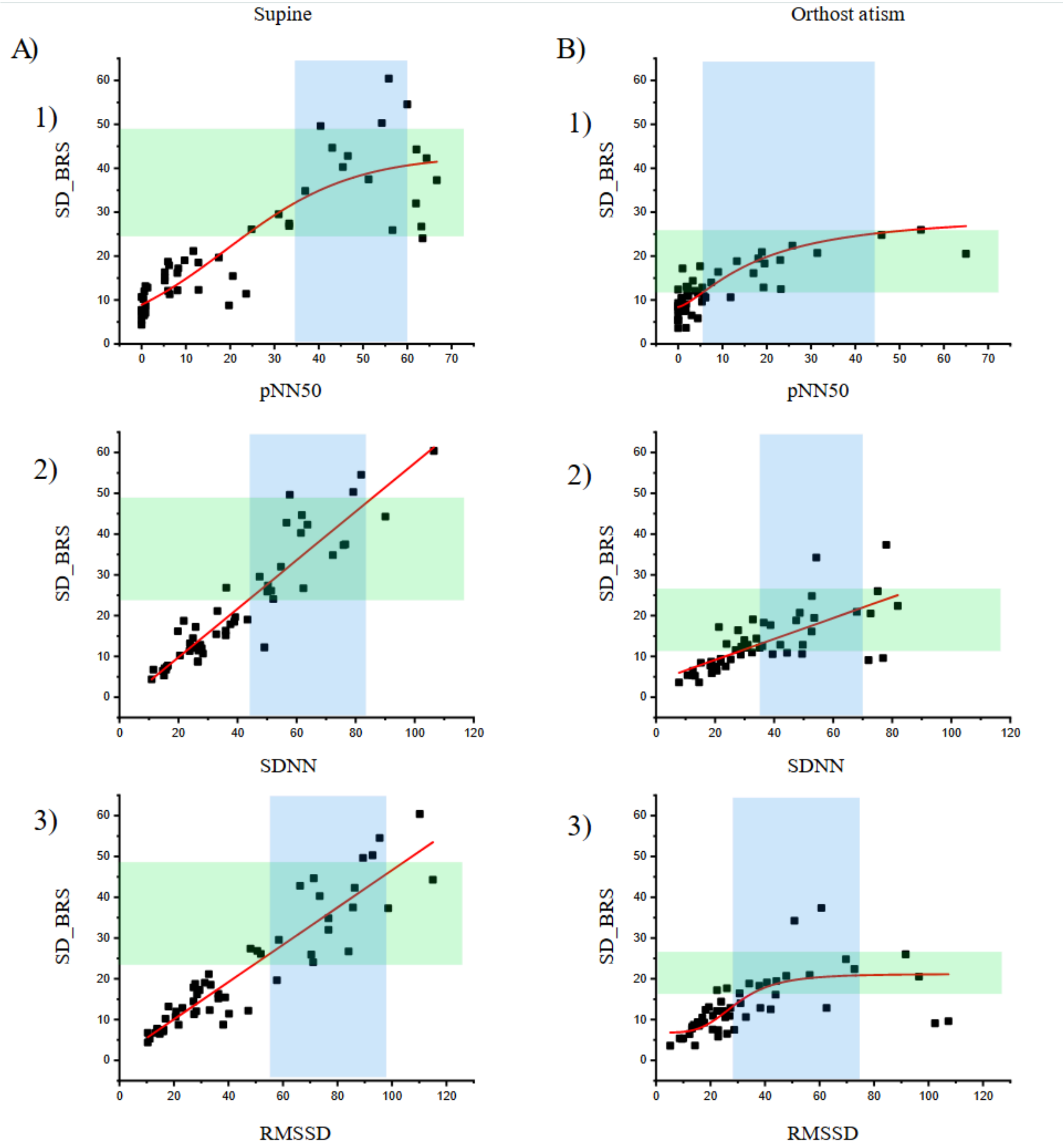


Fig 8

HRV variables have been placed on the X-axis to compare them with the SD BRS values obtained, which have been placed on the Y-axis in a scatter plot. Additionally, the range of the control group has been highlighted with a blue and green rectangle A1) Scatter plot of the pNN50 and SD BRS values during the supine position with an adjusted S-Logistic function. A2) Scatter plot of the SDNN and SD BRS values during the supine position with an adjusted linear function. A3) Scatter plot of the RMSSD and SD BRS values during the supine position with an adjusted linear function. B1) Scatter plot of the pNN50 and SD BRS values during the upright position with an adjusted Logistic function. B2) Scatter plot of the SDNN and SD BRS values during the upright position with an adjusted linear function. B3) Scatter plot of the RMSSD and SD BRS values during the upright position with an adjusted Logistic function.

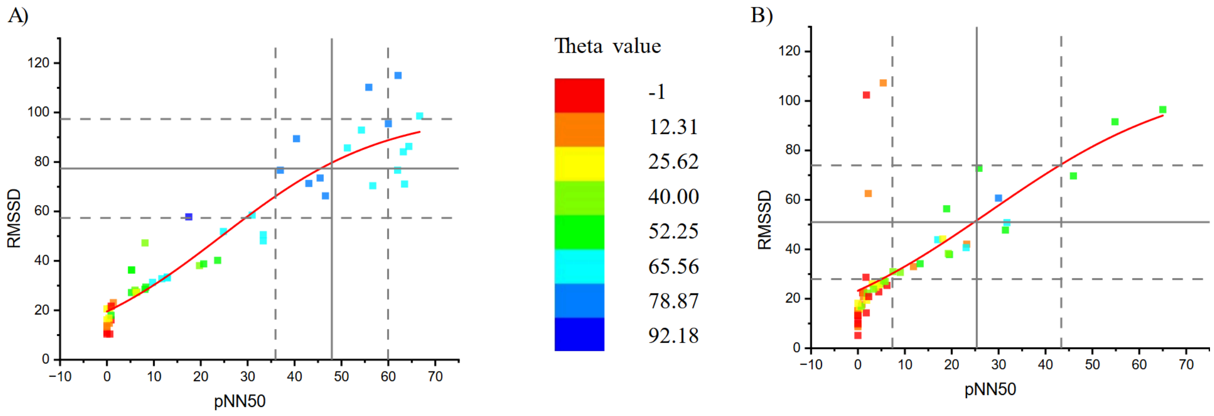


Fig 9

A) Scatter plot where pNN50 supine for all groups has been placed over the X-axis and the corresponding RMSSD supine value has been placed in the Y-axis.B) Scatter plot where pNN50 for orthostatism of all groups has been placed over the X-axis and the corresponding RMSSD for orthostatism value has been placed in the Y-axis. A single color gradient has been applied on both graphs.

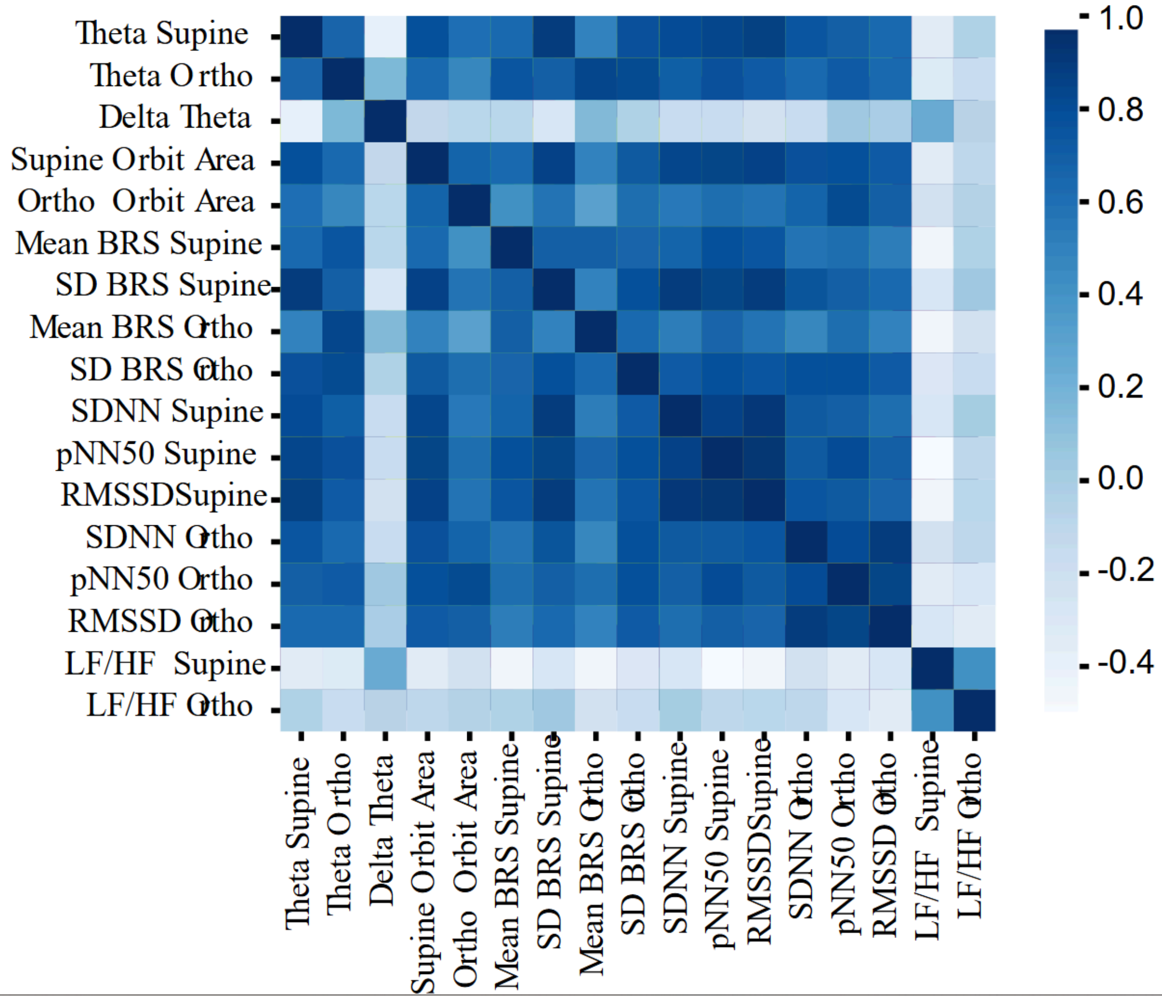


Fig 10.
Matrix with the canonical HRV and proposed Theta and SD BRS values during the supine and upright positions, where a heatmap was applied to match the correlation values obtained using the Spearman test.
