## Supplementary data for "A new method to measure Baroreflex sensitivity impairment in Long Covid patients with Hyperadrenergic POTS-like symptoms"

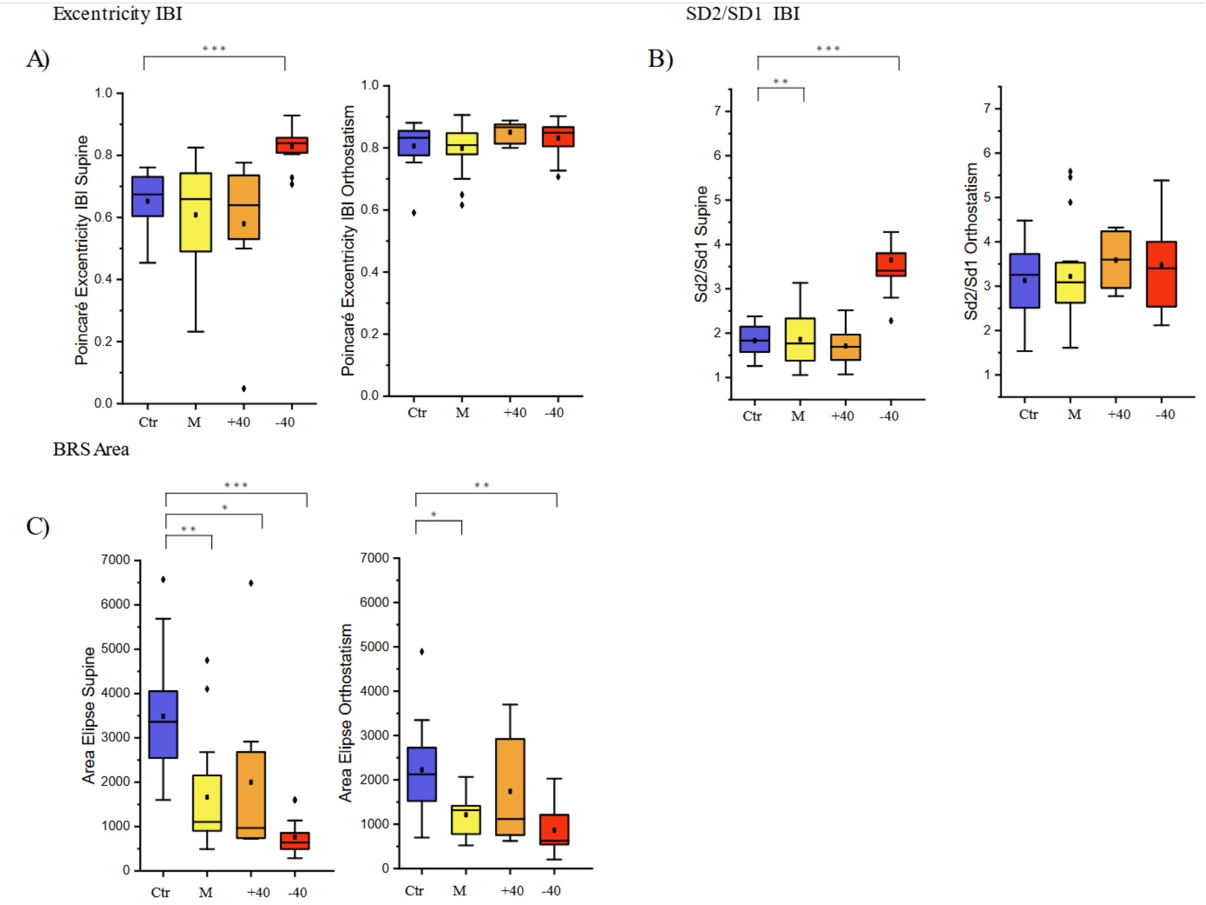


**Supplementary Fig 1.** Box plots with the data from Poincaré plots of each patient, such as eccentricity(A) and Sd2/Sd1(B), additionally the plot for the Area of the Theta ellipse(C) is also shown. To the indicate statistical significance with respect to the control group, an asterisk [*] was placed for a p<0.05, two asterisks [**] for p<0.001 and tree asterisks [***] for p<0.0001.

We also performed poincaré plots for the IBI values of all patients in order to observe their dispersion in continuous beat to beat data in which eccentricity was measured (Supplementary Fig 1.) for the supine and orthostatic poincaré graphs, where it was observed that the MP θ<40 group had a higher eccentricity value during the Supine period (0.82±0.05) which was significantly different than the other groups control(0.65±0.09; p<0.0001), MP-S(0.60±0.17; p<0.0001) & MP Δθ>40(0.57±0.23; p=0.000439). Although during the Orthostatic period all groups were not significantly different among them.

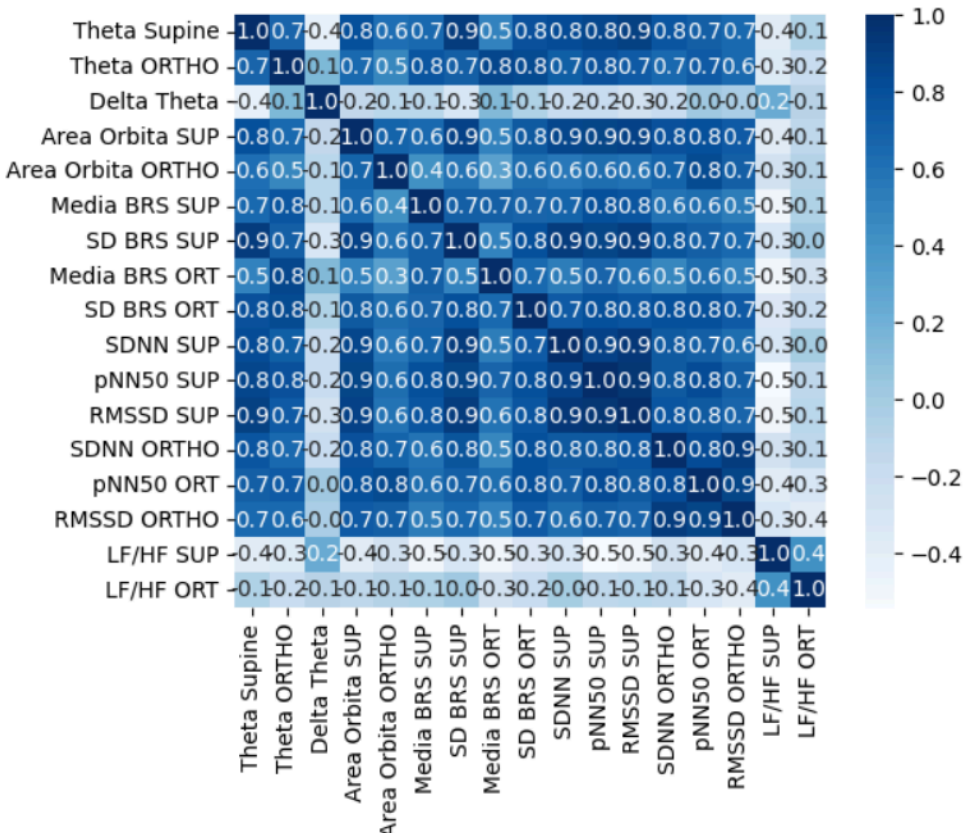


**Supplementary Fig. 2** Matrix with the canonical HRV and proposed Theta and SD BRS values during the supine and upright positions, where a heatmap was applied to match the correlation values obtained using the Spearman test.

**Comparison between groups:**

For the SBP during the supine and orthostatic periods, the patient groups did not show significant differences between them.

For the HR during the supine period, patients in the MP θ<40(76.25±6.55) group had a significantly different HR value than those the MP-S group (68.17±8.55; p=0.00647); during the Orthostatic period, the groups were not statistically different from each other.

**BRS Theta[θ]**

For the supine Theta value, the MP-S and MP Δθ>40 groups showed significant differences with the MP θ<40 group, while they were not significantly different between them (MP-S: p<0.0001; MP Δθ>40: p=0.00151). During the orthostatic period, the MP-S and MP Δθ>40 groups are significantly different (p=0.000429), and the MP-S and MP θ<40 groups are also significantly different (p<0.0001), although the MP Δθ>40 and the MP θ<40 groups were not significantly different during this period.

This shows that the MP Δθ>40 classification seems to be a transition state between the MP-S and the MP θ<40.

For the $\Delta$SYS, none of the groups is significantly different, although for $\Delta$Hr, the MP $\Delta\theta>40$ group was significantly different from the MP-S group ($p=0.02971$).

### BRS Histogram

BRS Mean during the supine period, the MP $\theta<40$ group had significantly lower values than those of the MP-S ($p<0.0001$) and MP $\Delta\theta>40$ ($p=0.0077$) groups; however, during orthostatism the MP-S group was significantly different than the MP $\Delta\theta>40$ ($p=0.000675$) and the MP $\theta<40$($p<0.0001$) groups, while these last two mentioned groups are not significantly different between them.

During the supine period, the BRS SD values of the MP $\theta<40$ group were significantly different than those of the MP-S ($p=0.00016$) and MP $\Delta\theta>40$ ($p=0.00033$) groups, which is also true during orthostatism (MP-S($p<0.0001$) & MP $\Delta\theta>40$ ($p=0.0019$)).

### HRV Values:

SDNN was significantly different for the <40 group during the supine (MP-S ($p=0.00296$), MP $\Delta\theta>40$ ($p=0.00371$)) and orthostatic (MP-S($p=0.00032$), MP $\Delta\theta>40$ ($p=0.00568$)) periods.

RMSSD was significantly different in the supine position for the MP $\theta<40$ group (MP-S ($p<0.0001$) and MP $\Delta\theta>40$ group ($p=0.00015$)); nonetheless, during orthostatism only the MP $\theta<40$ and MP-S groups showed significant differences between them ($p=0.00295$).

Similarly, the pNN50 values in the supine period showed significant differences in the MP $\theta<40$ group with the MP-S ($p<0.0001$) and MP $\Delta\theta>40$ ($p=0.00029$) groups, although during orthostatism only the MP $\theta<40$ group and the MP-S group ($p=0.00091$) showed significant differences between them.
